# A metabolic constraint in the kynurenine pathway drives mucosal inflammation in IBD

**DOI:** 10.1101/2024.08.08.24311598

**Authors:** Lina Welz, Danielle MM Harris, Na-Mi Kim, Abrar I Alsaadi, Qicong Wu, Mhmd Oumari, Jan Taubenheim, Valery Volk, Graziella Credido, Eric Koncina, Pranab K Mukherjee, Florian Tran, Laura Katharina Sievers, Polychronis Pavlidis, Nick Powell, Florian Rieder, Elisabeth Letellier, Silvio Waschina, Christoph Kaleta, Friedrich Feuerhake, Bram Verstockt, Melanie R McReynolds, Philip Rosenstiel, Stefan Schreiber, Konrad Aden

## Abstract

Inflammatory bowel disease (IBD) is associated with perturbed metabolism of the essential amino acid tryptophan (Trp). Whether increased degradation of Trp directly fuels mucosal inflammation or acts as a compensatory attempt to restore cellular energy levels via *de-novo* nicotinamide adenine dinucleotide (NAD^+^) synthesis is not understood. Employing a systems medicine approach on longitudinal IBD therapy intervention cohorts and targeted screening in preclinical IBD models, we discover that steady increases in Trp levels upon therapy success coincide with a rewiring of metabolic processes within the kynurenine pathway (KP). In detail, we identify that Trp catabolism in IBD is metabolically constrained at the level of quinolinate phosphorybosyltransferase (QPRT), leading to accumulation of quinolinic acid (Quin) and a decrease of NAD^+^. We further demonstrate that Trp degradation along the KP occurs locally in the inflamed intestinal mucosa and critically depends on janus kinase / signal transducers and activators of transcription (JAK/STAT) signalling. Subsequently, knockdown of *QPRT in-vitro* induces NAD^+^ depletion and a pro-inflammatory state, which can largely be rescued by bypassing QPRT via other NAD^+^ precursors. We hence propose a model of impaired *de-novo* NAD^+^ synthesis from Trp in IBD. These findings point towards the replenishment of NAD^+^ precursors as a novel therapeutic pathway in IBD.

## Introduction

Cellular energy deficiency has long been postulated as a critical pathophysiologic event in the context of inflammatory bowel disease (IBD) ^1^. Impairment of cellular nutrient supply, such as deficiency of nicotinamide adenine dinucleotide (NAD^+^), affects mitochondrial function and can thereby have deleterious effects on the equilibrium in the intestinal mucosa^2,3^. Upon inflammation, several pathways can be employed to restore exhausted cellular NAD^+^ levels. These include i) *de-novo* synthesis from the essential amino acid tryptophan (Trp) via the kynurenine pathway (KP), ii) salvage synthesis from nicotinamide (NAM) via nicotinamide phosphoribosyltransferase (NAMPT) or from nicotinamide riboside (NR) via NR kinases, and iii) the Preiss-Handler pathway, which utilizes nicotinic acid (NA) as a substrate and is exerted via nicotinic acid phosphoribosyltransferase (NAPRT) ^4–8^.

Increased metabolism of Trp has is a key metabolic occurrence in various chronic inflammatory diseases, including IBD ^9–11^. In this context, Trp is not only used for protein synthesis, but also catabolized into several bioactive derivatives important for immune regulation, cellular energy metabolism and redox balance ^12^. Trp is degraded along three main metabolic routes, with the vast majority (ca. 90%) of turnover occurring along the KP, with indoleamine-2,3-dioxygenase 1 (IDO1) constituting the initial and rate limiting enzyme ^13^. Only minor amounts of Trp are converted along the serotonin and indole pathway. In IBD, excessive catabolism of Trp correlates with disease activity indices, the extent of endoscopic inflammation and biochemical inflammation markers ^14,15^. Similarly, impaired intestinal uptake of Trp initiates intestinal inflammation, which can be overcome by supplementation of NR or NAM as precursors of NAD^+^ via the salvage pathway ^16–18^.

However, little is known about how the molecular functions of KP intermediates might directly fuel inflammatory responses. Recently, xanthurenic acid and kynurenic acid, metabolites produced within the KP, have been shown to be decreased in IBD patients due to reduced conversion via adenosine deaminase acting on tRNA (AADAT) ^19^. Interestingly, the precursor of *de-novo* NAD^+^ synthesis, quinolinic acid (Quin) is increased in individuals with IBD relative to the healthy population ^20,21^. Furthermore, enzymatic conversion of Quin is impaired upon stimulation of macrophages with lipopolysaccharide (LPS), leading to increased Quin levels *in-vitro* ^21^. Considering the evidence, we hypothesized that reduced conversion of Quin into nicotinic acid mononucleotide (NAMN) and subsequent deficiency of mucosal NAD^+^ during active inflammation of IBD may result from a metabolic restraint within the KP.

Using longitudinal IBD cohorts and a systems medicine approach, we show that Trp catabolism upon intestinal inflammation is characterized by a metabolic constraint at the conversion of Quin into NAD^+^, which is conferred by decreased expression of quinolinate phosphorybosyltransferase (QPRT) in the inflamed mucosa. Rewiring of Trp metabolism in IBD coincides with alleviation of mucosal inflammation. We further identify janus kinase / signal transducers and activators of transcription (JAK/STAT) as the major upstream driver of mucosal Trp degradation. Treatment with JAK inhibitors not only inhibits cytokine-mediated Trp wasting, but also restores the metabolic build-up of Quin. Along the same lines, inducing the metabolic constraint at Quin by knockdown of *QPRT* promotes a pro-inflammatory phenotype *in-vitro*, which can largely be overcome by restoring NAD^+^ levels via supplementation with NR. Our data therefore indicate that targeted cellular restoration of NAD^+^ levels, either by improving *de-novo* NAD^+^ synthesis from Trp or by boosting other NAD^+^ precursor pathways, provides a novel therapeutic entry point for targeted treatment of IBD by restoring cellular energy deficiency.

## Results

### Longitudinal rewiring of Trp metabolism indicates therapeutic efficacy in IBD

We assessed changes in serum Trp levels via serial measurements over the course of one year to understand how the disease state or long-term disease control, defined as 12-month therapy persistence on a newly induced biologic treatment, influences long-term Trp trajectories. We assessed serum Trp concentrations in 181 IBD patients (Crohn’s disease (CD), n = 99; ulcerative colitis (UC), n = 81) (Table S1). Patients achieving long-term disease control (n = 120) exhibited stable and continuous rise of serum Trp levels over the entire time course of 52 weeks (Fig. 1A), which was independent of disease entity (UC vs. CD, Fig. 1B), suggesting that rewiring of Trp metabolism coincides with disease control in IBD (Fig. 1AB). To formally assess the fate of the metabolized Trp in the context of IBD therapy, we examined longitudinal changes of Trp and corresponding metabolites of the kynurenine and serotonin pathways, and several indolic compounds (Fig. S1) in IBD patients (n = 134; n = 52 CD and n = 82 UC) at baseline, week 2 and week 14 after induction with biologic therapy (Fig. 1C, Table S2). Patients with available endoscopic scores at week 14 were classified into remission and non-remission categories based on either endoscopic Mayo score (eMayo, less than 2) or simple endoscopic score of CD (SES-CD; less than 3); in total, 34 patients reached remission and 57 did not. We used principal component analysis to identify gross changes in Trp metabolism associated with endoscopic remission at week 14 of treatment (Fig. S2A) and observed that metabolites corresponding to PC2 showed significant differences in week 14 between remission and non-remission (Fig. S2A).

**Figure 1:**
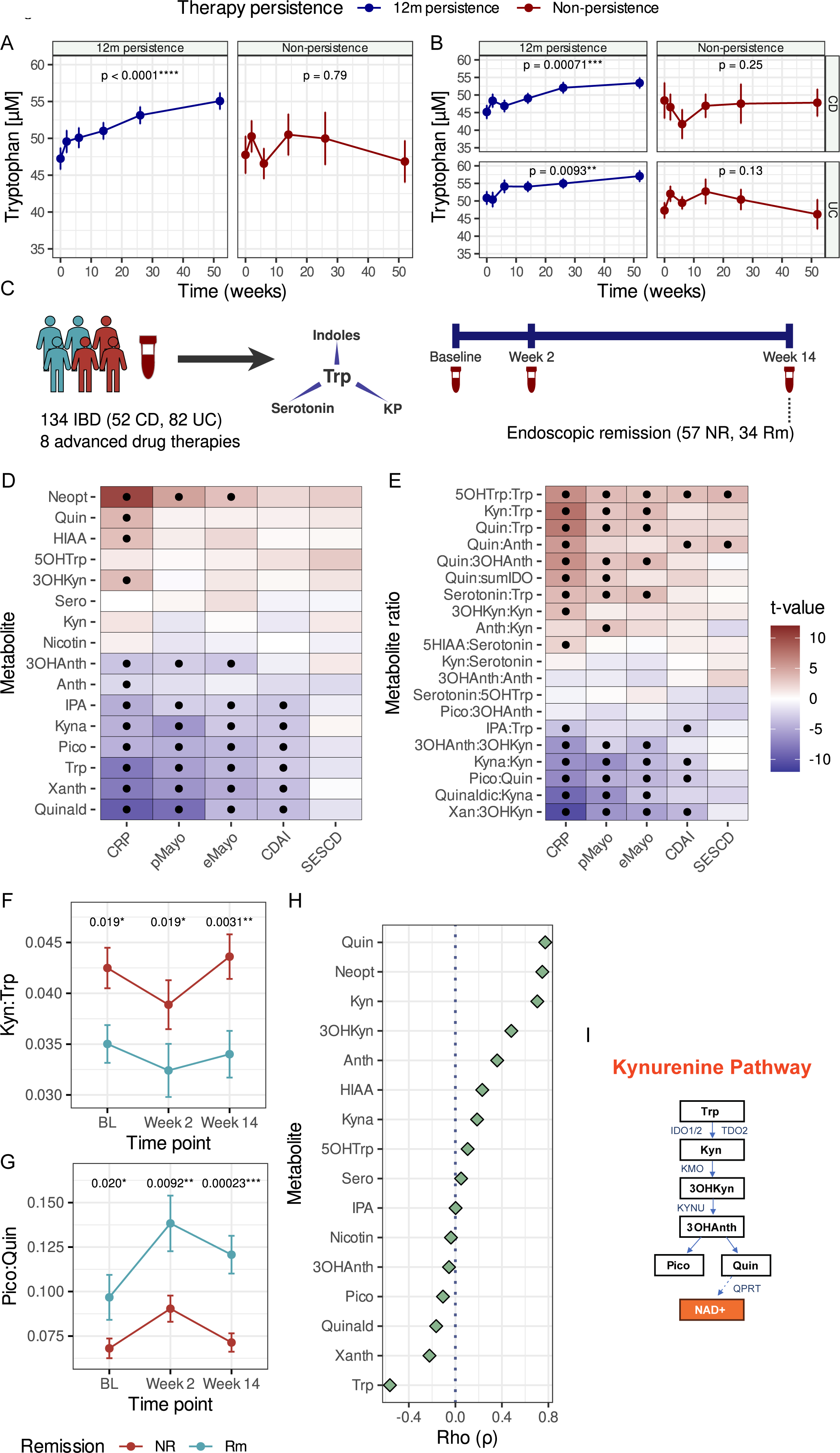
Longitudinal Trp (derivative) trajectories indicate restored Trp metabolism with therapy success. (A,B) Longitudinal trajectory of Trp for patients with IBD who persisted on their prescribed biologic for 12 months (12m persistence), or whose therapy was discontinued at some point within 12 months from baseline. All IBD patients were included together, and separately checked by diagnosis (UC/CD). Trp was measured using high performance liquid chromatography. (C) Cohort structure for the analysis of downstream Trp derivatives measured using targeted mass spectrometry. Endoscopic remission was determined at week 14 based on available endoscopic Mayo (< 2) or SES-CD (< 3). (D,E) Linear mixed models were used to identify links between the Trp derivatives measured and the indicated metrics of disease activity. Black dots indicate statistical significance at a level of p_adj_ = 0.05. All patients with available disease activity metrics were used for the analysis. (F,G) Selected trajectories from the analysis of the dynamics of Trp derivatives in patients who entered endoscopic remission by week 14. Linear mixed models were used to assess the difference, with contrast analysis applied to assess the group interaction at each time point. Only patients with endoscopic scores at week 14 were included. (H) Spearman correlation of baseline Trp derivatives with the Kyn:Trp ratio. (I) Simplified overview of the kynurenine pathway with specific emphasis on *de-novo* NAD^+^ synthesis. Neopt: Neopterin, Quin: Quinolinic acid, HIAA: 5-Hydroxy indoleacetic acid, 3OHKyn: 3-Hydroxy-kynurenine, 5OHTrp: 5-Hydroxy-tryptophan, Sero: Serotonin, Kyn: Kynurenine, Nicotin: Nicotinamide, Anth: Anthranilic acid, 3OHAnth: 3-OH-Anthranilic acid, IPA: Indole-3-propionic acid, Kyna: Kynurenic acid, Pico: Picolinic acid, Trp: Tryptophan, Xanth: Xanthurenic acid, Quinald: Quinaldehyde, Quin:sumIDO: the ratio of quinolinic acid to IDO-derived metabolites, calculated as Quinolinic.acid / (3-OH-Anthranilic acid + 3-OH-Kynurenine + Kynurenic acid + Kynurenine + Picolinic acid + Quinolinic acid + Xanthurenic acid). IDO1/2: Indoleamine 2,3-dioxygenase 1/2, KMO: Kynurenine-3-monooxygenase, KYNU: Kynureninase, NAD^+^: Nicotinamide adenine dinucleotide, TDO: Tryptophan-2,3-dioxygenase.

We next assessed individual associations between metabolites or metabolite ratios and clinical (Crohn’s disease activity index: CDAI, CD; partial Mayo: pMayo, UC), endoscopic (SES-CD, CD; eMayo, UC) and biochemical (C-reactive protein: CRP) disease activity indices (Fig. 1D,E). In line with previous studies ^19,20,22^, the kynurenine (Kyn) to Trp (Kyn:Trp) ratio was higher in individuals who would not enter remission at week 14 at weeks 0, 2 and 14 (Fig. 1F). Surprisingly, we found that the ratio of downstream metabolites picolinic acid (Pic) and Quin (Pico:Quin), also exhibited differences at each time point tested: individuals who would enter remission by week 14 had significantly higher Pico:Quin (Fig. 1G). In addition, the correlation between Quin and Kyn:Trp was stronger for Quin (Spearman’s rho, ρ = 0.77, FDR < 0.0001) compared to Pico (ρ = −0.11, FDR = 0.38) (Fig. 1H). We further ruled out that these differences simply resulted from drastic contrasts in baseline inflammation levels between remission and non-remission (Fig. S2, p_CRP_ = 0.19, p_CDAI_ = 0.11, p_tMayo_ = 0.51). These data indicate that the direction through which Trp is degraded along the KP (Fig. 1I) associates with disease activity and therapy response in IBD, and in particular, imply a potential role of Quin accumulation in the IBD disease course.

### Transcriptome-aided metabolic modelling reveals reduced Quin to NAD^+^ occurs in the mucosa

We next aimed to disentangle the local and systemic trajectories of Trp degradation in IBD in relation to disease activity. To this purpose, we took advantage of a longitudinal clinical cohort of IBD patients (n = 62; UC, n = 30; CD, n = 32), in which serial biosampling of blood and intestinal biopsies was conducted before and at indicated time points after induction with biologic therapy (Fig. 2A, Table S3) ^23–25^. By estimating the reaction activity scores of known enzymatic reactions involved in Trp catabolizing in bulk transcriptome data derived from blood and intestinal mucosa ^26,27^, we aimed to understand i) which catabolic events of Trp degradation are most predominantly activated during intestinal inflammation and ii) at which anatomical location (blood vs. intestine) those events occur.

**Figure 2:**
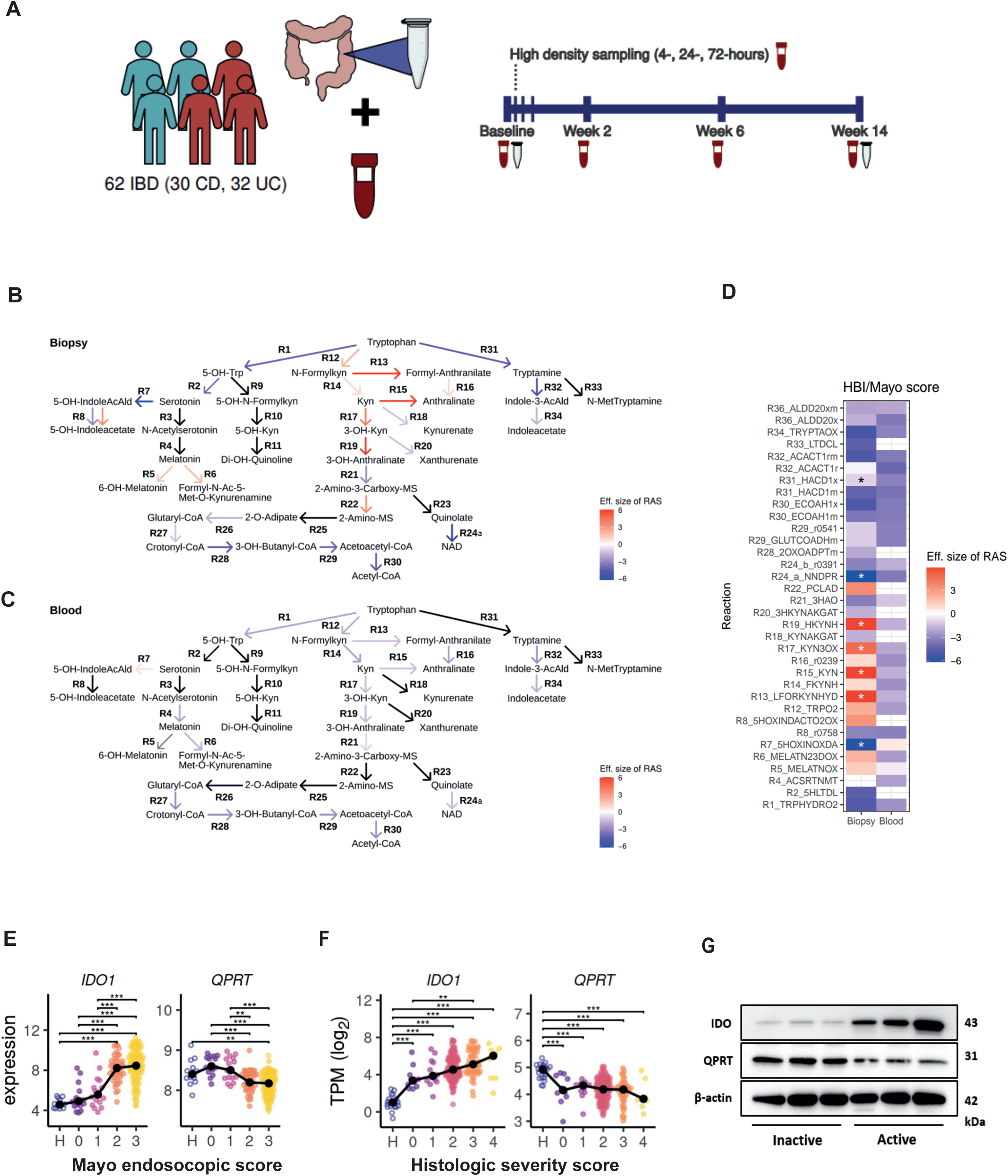
Transcriptome-aided metabolic modelling reveals mucosal roadblock at conversion of Quin into NAD^+^. (A) Cohort overview: Whole blood and biopsies were obtained within two longitudinal IBD intervention cohorts sampled in northern Germany (n = 62 IBD, n = 32 UC, n = 30 CD) over a period of 14 weeks at the indicated timepoints. Patients were treatment-naïve for biologic treatment and were introduced to either anti-TNFα (n = 22), anti-α4β7-Integrin (n = 21), anti-IL6-trans (n = 16) or anti-IL6-R (n = 3). Patients were clinically and endoscopically monitored. (B,C) Expression of genes involved in the degradation of Trp was associated with disease activity indices (HBI, CD; total Mayo, UC). The human metabolic model (recon3D) was used to extract the Trp degradation pathway from it. Gene expression values were mapped to Trp degrading pathways by applying the gene-protein-reaction relations of recon3D (github.com/Porthmeus/CORPSE) to estimate reaction activity scores (RAS). RAS were then associated with HBI/Mayo scores in linear mixed models; the effect size (t-value) of the linear mixed model was color-coded in the arrows displaying the reactions facilitated by the enzymes. Black arrows indicate missing expression data due to pre-filtering or missing gene-to-reaction rule. All p-values were adjusted with the Benjamini-Hochberger correction. (D) Enzyme expression for the degradation of Trp was associated with disease activity (HBI/Mayo) and the effect size (t-value) of the linear mixed model were displayed as a heatmap. Significant changes occur in: R7, R13, R15, R19, R24a, R32 (FDR p < 0.05). (E,F) Expression of *IDO1* and *QPRT* in association with i) endoscopic IBD disease activity in 166 biopsies from 67 UC patients (treated with either infliximab or vedolizumab) and 12 samples from 12 healthy donors (dataset GSE73661 ^30^), ii) the histology severity score (HSS) of 206 new-onset UC patients and 20 healthy controls (dataset GSE109142 ^29^). Statistical analysis was performed with a one-way-ANOVA followed by Tukey’s HSD post-hoc test. ∗∗*P* < 0.01; ∗∗∗*P* < 0.001. Black dots and connecting lines show the median expression. (G) Protein levels of IDO and QPRT were assessed in human mucosal biopsies (inactive (eMayo / Nancy 0) and active (eMayo 3 / Nancy 4), n = 3 biological replicates / group), ß-Actin served as a housekeeper (representative of n = 1 individual experiment). IDO1: Indoleamine 2,3-dioxygenase 1, QPRT: Quinolinate phosphorybosyltransferase.

We first correlated reaction abundance of Trp degrading pathways with clinical disease activity indices (Harvey-Bradshaw Index (HBI), CD; total Mayo score, UC). Significant correlations of Trp degrading enzymes with disease activity were noted in the mucosa, but not in blood (Fig. 2B-D). We concluded that the inflamed mucosa represents the primary anatomical compartment of Trp degradation during intestinal inflammation. We next inferred the flux of Trp metabolism in the inflamed mucosa and observed a decreased reaction abundance for mucosal genes involved in the conversion of Quin into nicotinic acid mononucleotide (NAMN) (reaction ID R24a), indicating that accumulation of Quin in the serum metabolome might be the result of insufficient enzymatic conversion into NAD^+^ (Fig. 2B,D).

To confirm this hypothesis, we assessed the expression of quinolinate phosphorybosyltransferase (*QPRT*), which converts Quin into NAMN and is the final unique enzyme before *de-novo* NAD^+^ synthesis merges with the Preiss-Handler pathway ^28^, as well as the expression of *IDO1* (indoleamine 2,3-dioxygenase 1), the enzyme that shuttles Trp into the KP. Mucosal expressions of *IDO1* and *QPRT* in publicly available transcriptome datasets from sigmoid biopsies from UC patients significantly correlated with both endoscopic (Fig. 2E) and histologic (Fig. 2F) disease activity ^29,30^. Whereas *IDO1* expression increased with inflammatory severity, *QPRT* levels decreased, again implying a metabolic constraint in turning over the increased moieties of Quin that result from overactivation of the KP into NAD^+^ (Fig. 2E,F). To confirm increased protein expression of IDO1 and reduced abundance of QPRT with increasing mucosal inflammation, we used protein lysates of intestinal biopsies derived from UC patients (active defined as eMayo = 3; inactive defined as eMayo = 0) for a western blot, which confirmed a positive correlation of IDO1 and negative correlation of QPRT protein expression in non-inflamed vs. inflamed colon tissue, respectively (Fig. 2G).

### DSS colitis confirms mucosal metabolic constraint at Quin

We next used the preclinical dextran sodium sulfate (DSS) colitis model to conduct targeted metabolomics of i) serum samples that were collected every other day as well as ii) tissues from different states of intestinal inflammation (*flare-up* (day 5) vs. *flare* (day 8/11)) (Fig. 3A). Beginning from day 3 (d3) of the DSS colitis, we observed an increase in serum Kyn which resulted in heightened Kyn:Trp serum ratios from d5-d11 as measured by LC-MS (liquid chromatography mass spectrometry) (Fig. S5). No further significant inflammation-associated changes of Trp metabolites were observed in murine serum, although Quin tended to increase with colitis severity (Fig. S5). However, when systematically analysing the levels of Trp and downstream metabolites of the KP in all anatomical locations *post-mortem*, we found that Trp degradation predominantly occurs at the site of inflammation, namely the colon (Fig. 3B,D). This implies that the observed accumulation of metabolites of the KP in serum of IBD patients might be majorly driven by mucosal Trp degradation. In addition, we confirmed that catabolism of Trp led to an accumulation of mucosal Quin, whereas NAD(H) levels were significantly reduced (Fig. 3F-H). Hence, our data corroborate the findings of mucosal accumulation of Quin and a corresponding decrease of NAD(H) levels as critical metabolic events in mucosal inflammation.

**Figure 3:**
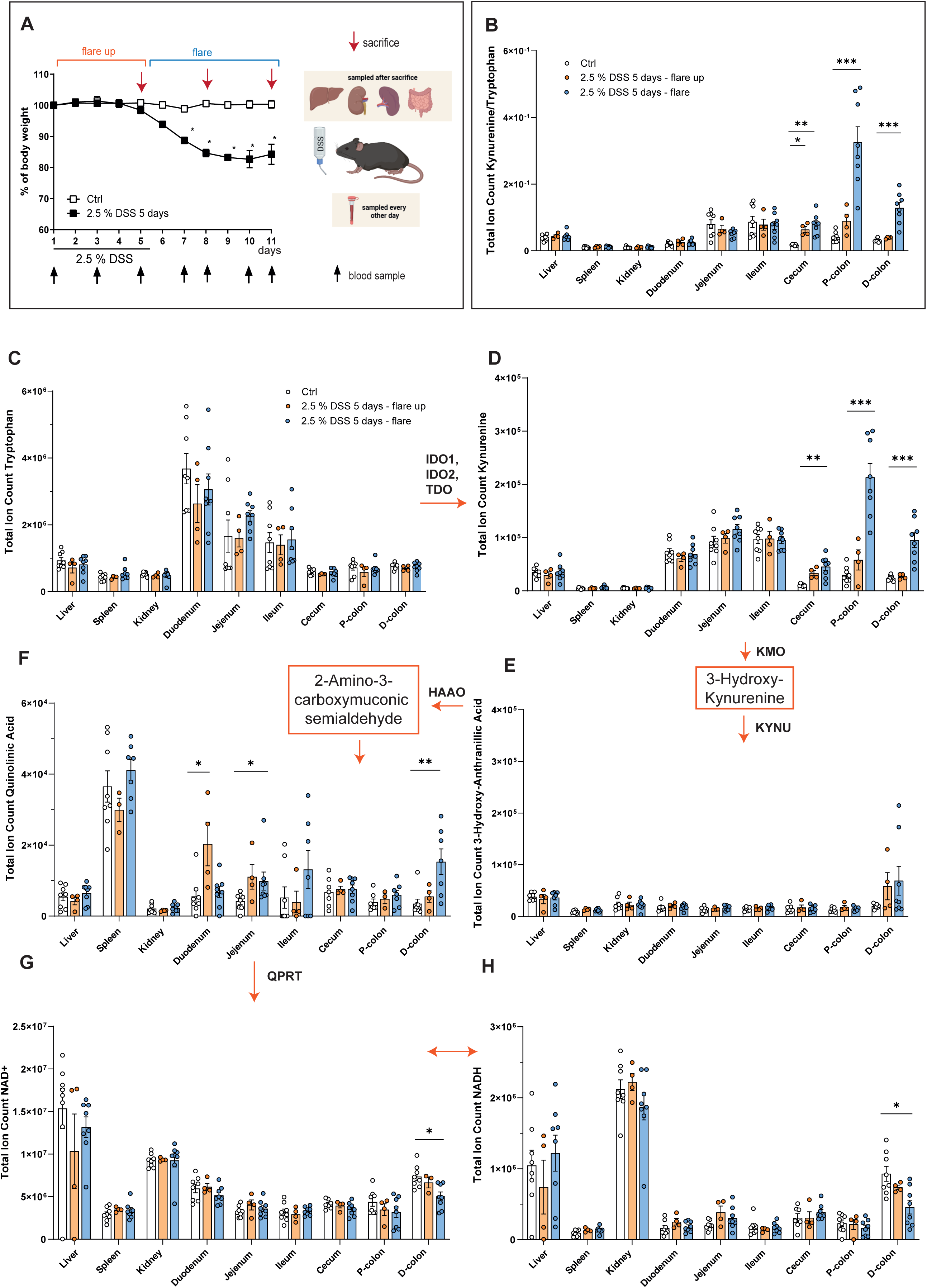
DSS colitis validates mucosal metabolic constraint at Quin. (A) Experimental set-up: 8-12 weeks old, male C57BL/6J mice (n = 20) were supplied with drinking water (white, control) or 2.5% DSS for 5 days (black, normal drinking water was supplied from day 6 on). Blood was collected every over day (indicated by black arrows). Animals were euthanized (each n = 4 / control or DSS group) on day 5, day 8 and day 11 (indicated by red arrows). (B) Kyn/Trp ratio of the metabolites measured in (C) and (D). (C-H) Total ion counts of Trp metabolites were measured in the indicated tissues by LC-MS. Groups: <u>White</u> = control groups of mice sacrificed on days 5 and 8 (n = 8); <u>orange</u>: DSS colitis flare-up (sacrifice on day 5, n = 4), <u>blue</u>: DSS colitis flare (sacrifice on day 8, n = 4 and sacrifice on day 11, n = 4). Each dot represents the measurements of one individual mouse. All data are presented as mean ± SEM. Statistical analysis was performed with Kruskal-Wallis test. ∗*P* < 0.05; ∗∗*P* < 0.01; ∗∗∗*P* < 0.001; ∗∗∗∗*P* < 0.0001. D-Colon: Distal colon, HAAO: 3-Hydroxyanthranilate 3,4-dioxygenase, IDO1: Indoleamine 2,3-dioxygenase 1, KMO: Kynurenine-3-monooxygenase, Kyn: Kynurenine, KYNU: Kynureninase, NAD^+^/NADH: Nicotinamide adenine dinucleotide, P-Colon: Proximal colon, QPRT: Quinolinate phosphorybosyltransferase, TDO: Tryptophan-2,3-dioxygenase, Trp: Tryptophan.

### Mucosal Trp degradation in IBD is JAK/STAT-dependent

We next aimed to identify upstream transcriptional signatures that causally drive Trp wasting in the intestinal mucosa. Employing an integrative approach, we combined bulk transcriptome analysis of peripheral blood and intestinal biopsies with serum metabolomics (longitudinal sampling from UC, n = 30; CD, n = 32; Fig. 2A) to integrate gene expression of Trp degrading genes with serum Trp catabolism. We aimed to identify commonly regulated signal transduction pathways that correlate with i) serum Kyn:Trp ratios and ii) disease activity scores (HBI / total Mayo score) using linear mixed models.

Among the signal transduction pathways found for both approaches, we identified two pathways, nuclear factor ‘kappa-light-chain-enhancer’ of activated B-cells (NF-κB) and janus kinases / signal transducer and activator of transcription proteins (JAK/STAT) as upregulated in inflamed tissue (Fig. 4A,B). When assessing overlapping signalling transduction pathways between blood and tissue in association with disease activity scores (HBI / total Mayo) or serum Kyn:Trp ratios, we validated the JAK/STAT pathway as upregulated in tissue upon high disease activity and Kyn:Trp ratios (Fig S6A,B).

**Figure 4:**
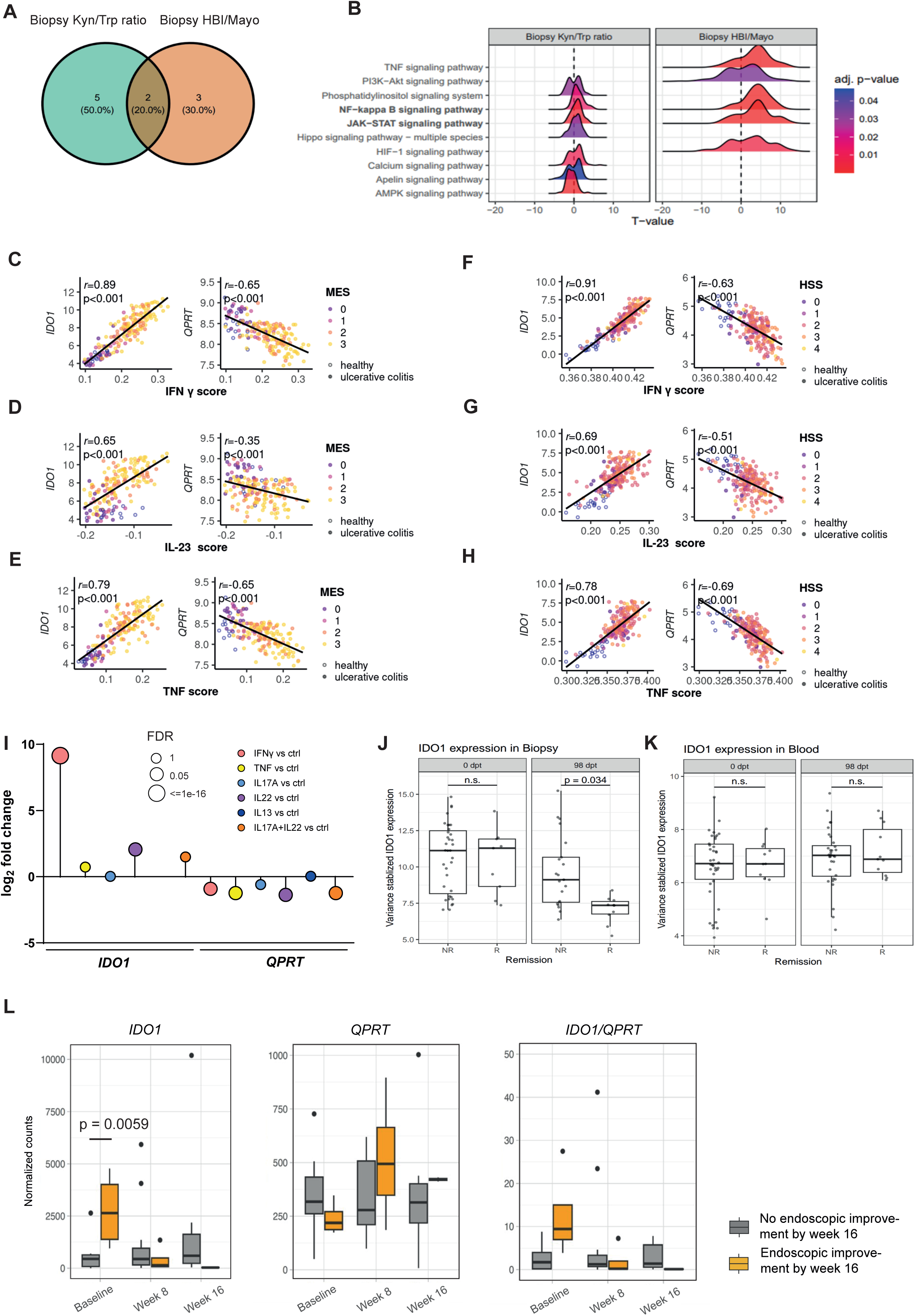
Mucosal Trp degradation in IBD is JAK/STAT-dependent. (A,B) Whole blood and biopsies were obtained within two longitudinal IBD intervention cohorts sampled in northern Germany (n = 62 IBD, n = 32 UC, n = 30 CD) over a period of 14 weeks at the indicated timepoints. Patients were treatment-naïve for biologic treatment and were introduced to either anti-TNFα (n = 22), anti-α4β7-Integrin (n = 21), anti-IL6-trans (n = 16) or anti-IL6-R (n = 3). Transcriptomics were conducted on whole blood and biopsies, and serum samples were used for metabolomics. Differential gene expression analysis was performed using linear mixed models to associate changes in gene expression with changes in disease activity (HBI/Mayo) and Kyn/Trp ratios in serum. Afterwards, we employed gene set enrichment (GSEA) and hypergeometric tests on the signal transduction pathways annotated by KEGG. All p-values adjusted with the Benjamini-Hochberger correction. (C-H) For the analyses performed in Fig. 2E and F, the putative upstream cytokine network (TNFα, IFNγ and IL-23) was inferred. Pearson’s correlation coefficients r and p-values are shown. (I) Gene expression was assessed in the RNA sequencing dataset from human colonic organoids and corresponding analysis from Pavlidis et al. using the same varying intercepts hierarchical modelling framework described therein ^61^. *IDO1* and *QPRT* expression were assessed in the transcriptomes of colonoids from HC after treatment with the following cytokines: IFNγ, IL-17A, IL-22, TNFα (each n = 4 biological replicates) and IL-13 (n = 3 biological replicates). (J,K) *IDO1* expression was found to be one of the top three genes differentially expressed in tissue between CD and UC remitters and non-remitters in week 14 (defined as a HBI <5 or a partial Mayo score <2) of the cohort used for the analysis performed in Fig. 4A,B (baseline: 0 dpt, week 14: 98 dpt; remitter: baseline −9 biopsies and 11 blood samples; week 14 - 10 biopsies and 11 blood samples; non-remitter: baseline - 35 biopsies and 38 blood samples; week 14 - 21 biopsies and 32 blood samples). (L) Normalized counts of *IDO1* and *QPRT* expression and *IDO1*/*QPRT* ratio were compared between endoscopic responders and non-responders in mucosal biopsies of UC patients treated with Tofacitinib (4 responders, 11 non-responders). Endoscopic improvement was defined as a Mayo endoscopic subscore ≤1 ^62^. Statistical analysis was performed using Mann-Whitney-U test. Dpt: Days post treatment initiation, FDR: False discovery rate, IDO1: Indoleamine 2,3-dioxygenase 1, Kyn: Kynurenine, QPRT: Quinolinate phosphorybosyltransferase, Trp: Tryptophan.

We further confirmed the molecular link between JAK/STAT signalling and mucosal expression of the key KP enzymes *IDO1,* a known JAK/STAT downstream target gene ^31–33^, and *QPRT* by inferring the putative upstream cytokine network ^34^. We found that mucosal *IDO1* induction and *QPRT* downregulation significantly correlated with IFNγ, IL-23 and to a lesser extent with TNF, implying that mucosal upstream cytokines signalling via JAK/STAT crucially drive Trp breakdown and Quin accumulation via the KP (Fig. 4C-H). Importantly, the JAK/STAT dependency of increased *IDO1* gene expression and reduced *QPRT* gene expression could be validated in the epithelial compartment upon stimulation of human colonic organoids with different JAK/STAT inducing pro-inflammatory cytokines (Fig. 4I). Lastly, we assessed the differential expression of mucosal genes (baseline vs. week 14) in IBD patients achieving clinical remission at week 14 and identified in total 3 genes significantly downregulated at week 14, namely regenerating family member 1 alpha (*REG1A*), FAM3 metabolism regulating signalling Molecule B (*FAM3B*) (data not shown) and *IDO1* (Fig. 4J,K). Interestingly, *IDO1* was not differentially expressed in the blood, which is in line with our observation of tissue-specific regulation of Trp metabolism (Fig. 4K). Hence, we concluded that amelioration of JAK/STAT-driven Trp degradation might be a crucial pathophysiological event in the manifestation of clinical remission in IBD. We therefore independently validated IDO1 as a putative target of JAK inhibition in IBD and assessed the mucosal transcriptome data of UC patients treated with the JAK inhibitor tofacitinib ^35^. *IDO1* expression was significantly upregulated and *QPRT* downregulated at baseline between endoscopic responders and non-responders (Fig. 4L), which was not observed for UC patients treated with infliximab or vedolizumab (Fig. S6C,D), indicating that baseline IDO1 expression might be a potential biomarker for therapy response to JAK inhibitor treatment.

### Single cell sequencing of intestinal mucosa reveals distinct cellular subsets involved in Trp catabolism

We further tried to delineate the specific cell types that are involved in driving Trp degradation in the intestinal wall of IBD patients. We therefore employed scRNA sequencing data from intestinal tissue of healthy control HC (n=7) and CD patients (n=13, inflamed vs. non-inflamed) for assessing the expression of *IDO1* and *QPRT* ^36^.

We surprisingly identified two distinct cellular clusters of *IDO1-* and *QPRT-* expressing cells with no overlap of cells co-expressing *IDO1* and *QPRT* (Fig. 5A-E). We found *IDO1* expression almost exclusively confined to myeloid cells, suggesting that the rate limiting conversion of Trp into Kyn occurs mostly in the myeloid compartment (Fig. 5A,C,F). In contrast, expression of *QPRT*, needed for the conversion of Quin into NAD^+^, was most strongly observed in fibroblasts and epithelial cells (Fig. 5A,B,F).

**Figure 5:**
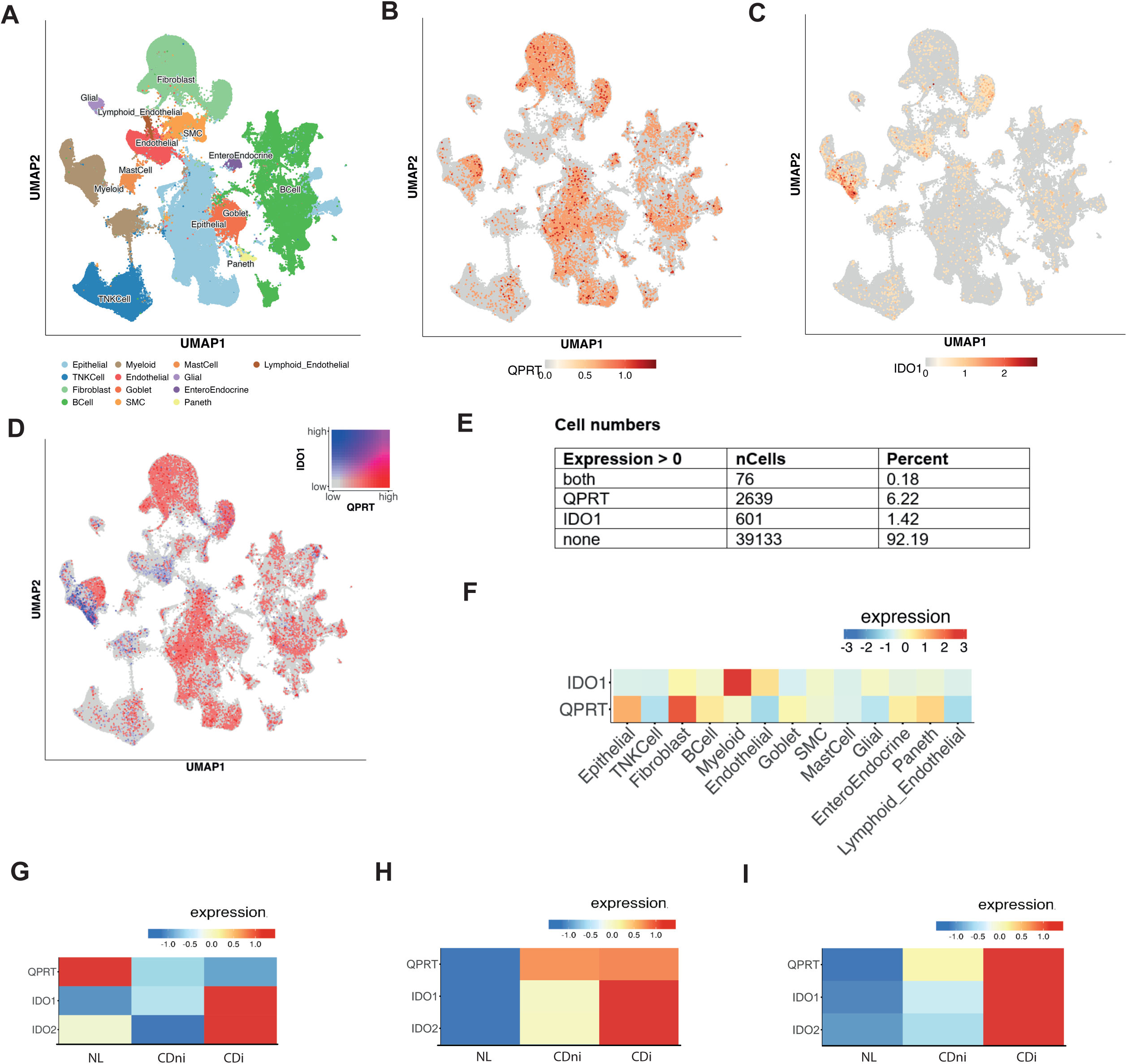
Single cell sequencing of intestinal mucosa reveals cellular subsets involved in Trp catabolism. Single cell transcriptional analysis of expression of *QPRT* and *IDO1* in intestinal tissues obtained from Crohn’s disease (CD, n = 13) or non-IBD (normal, “NL”, n = 7) patients. (A) Uniform Manifold Approximation and Projection (UMAP) showing the cell types identified in the single cell dataset with (B) *QPRT and* (C) *IDO1* expression. (D, E) Expression of both *IDO1* and *QPRT* across different cell types was quantified (F) Heatmap depicting expression of *QPRT* and *IDO1* in intestinal cell sub-types. (G-I) Expression of *QPRT*, *IDO1* and *IDO2* in (G) fibroblasts, (H) myeloid cells and (I) epithelial cells across the intestinal biopsies of non-diseased (NL), non-inflamed CD (CDni) and inflamed CD (CDi) patients’ phenotypes. Data sets and analysis were based on RNA scSeq data generated in a prior publication ^36^.

We next questioned in which cellular compartment the metabolic constraint at *QPRT* identified in bulk transcriptomics occurs. Based on the study from Minhas et al. ^21^demonstrating the Quin-metabolic constraint in macrophages, we expected to find a downregulation of *QPRT* expression with increasing disease activity especially in the myeloid compartment (Fig. 5H). However, a disease activity-dependent *QPRT* reduction was only observed in fibroblasts (Fig. 5G-I). We therefore concluded that the metabolic constraint at the level of QPRT originates from both - the loss of *QPRT*-expressing cells in the context of active inflammation (i.e., loss of the intestinal epithelium) and the inability of cell types (i.e., fibroblast) to adequately express *QPRT* in response to higher NAD^+^ demand.

### JAK inhibition restores IFNγ-driven Trp conversion via IDO1

We next intended to confirm the JAK-dependency of *IDO1* in myeloid cells. As increased Kyn:Trp ratio approximates IDO1 activity and our analysis revealed a dependence on JAK/STAT signalling (Fig. 4), we tested the hypothesis that JAK inhibition abrogates Trp degradation in the myeloid compartment. Embarking on previous results of human immune cells exerting a critical role in Trp degradation along the KP in inflammation ^21^, we treated human PBMC with lipopolysaccharide (LPS) to induce a pro-inflammatory state. LPS stimulation significantly upregulated *IDO1,* but not *IDO2 or* tryptophan-2,3-dioxygenase (*TDO*) expression (Fig. 6A, Fig. S7A,B). Correspondingly, LC-MS analysis of cell lysates and supernatants following LPS treatment revealed a significantly elevated Kyn:Trp ratio indicative of LPS-induced Trp degradation (Fig. S7C-H). To uncover the cell signalling pathways involved, we analyzed gene expression of immune cell-specific cytokines known to signal via JAK/STAT after LPS treatment in human PBMC (including IFNγ, TNFα, IL-2, IL-6, IL-10, IL-12, IL-22 and IL-23) (Fig. 6B). Of the cytokines tested, only IFNγ had a significant impact on *IDO1* and *IDO2* gene expression and was therefore used for further analysis (Fig. 6C,D).

**Figure 6:**
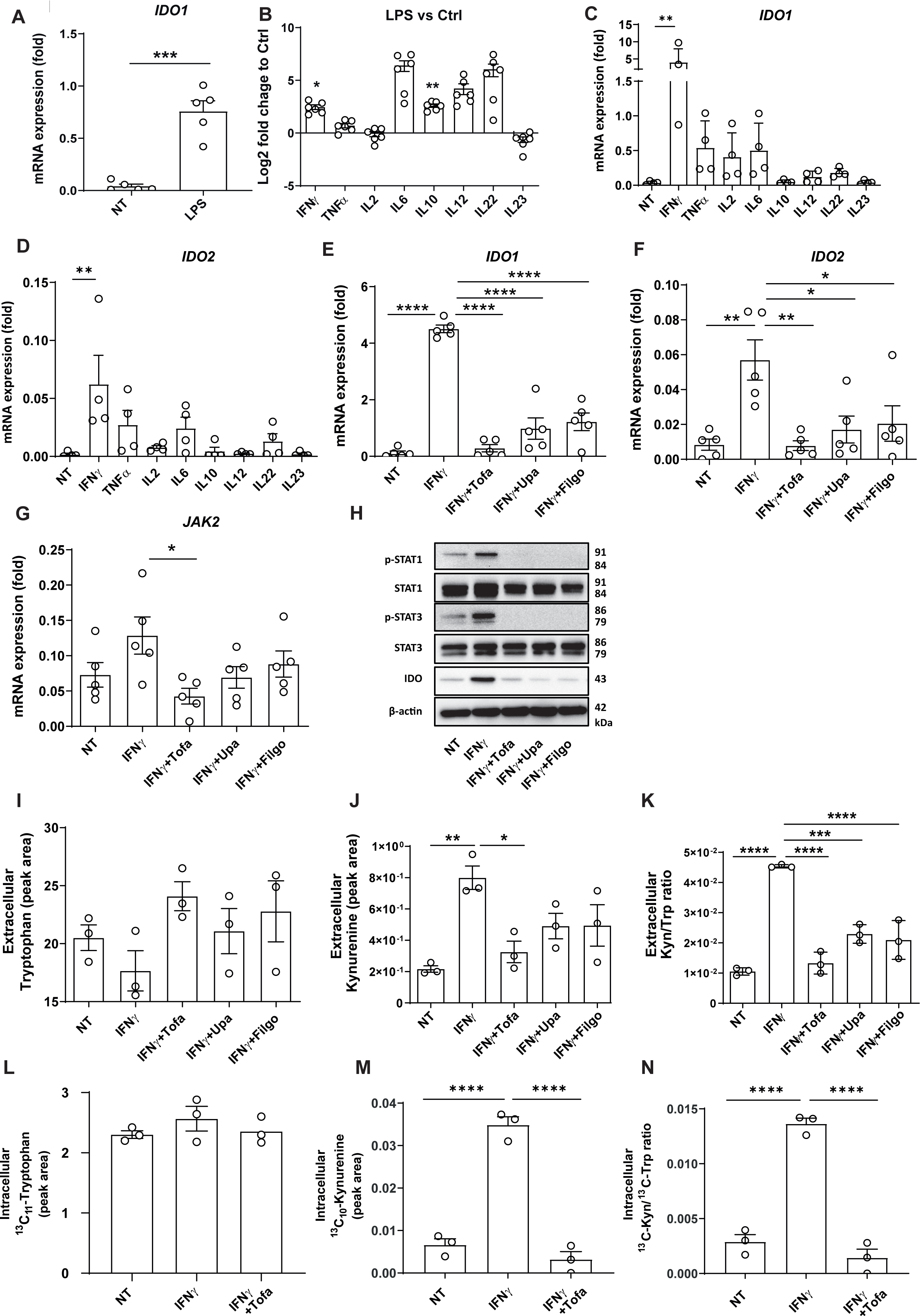
JAK inhibition restores IFNγ-driven Trp conversion via IDO1. (A,B) Human PBMC were treated with LPS (100 ng/ml) for 24 hours. (A) Relative gene expression was assessed by RT-qPCR (n = 2 technical replicates, representative of n = 5 individual experiments). (B) Relative gene expression levels were expressed as log2 fold changes (n = 2 technical replicates, representative of n = 6 individual experiments). (C,D Relative gene expression in human PBMC was detected after 24 hours of stimulation with 10 ng/ml of the named pro- or anti- inflammatory cytokines by RT-qPCR (n = 2 technical replicates, representative of n = 4 individual experiments). (E-K) Human PBMC were incubated with IFNγ (10 ng/ml) ±1 µM tofacitinib (Tofa), 0.1 µM upadacitinib (Upa) or 10 µM filgotinib (Filgo) for 24 hours. (E-G) Relative gene expression levels were measured by RT-qPCR (n = 2 technical replicates, representative of n = 5 individual experiments). For gene expression analyses, ß2- Mikroglobulin was used as housekeeping gene. (H) Protein levels of (phospho)STAT1 and (phospho)STAT3 as well as IDO1 were detected by WB and normalized to ß-Actin (representative of n = 3 individual experiments). (I-K) Extracellular Trp and Kyn levels were measured by LC-MS (n = 3 technical replicates, representative of n = 3 individual experiments; intracellular metabolites of this experimental set-up can be found in Fig. S7I-K). (L-N) Human PBMC were incubated with IFNγ (10 ng/ml) ± Tofa (1 µM) for 24 hours. Intracellular levels of ^13^C_11_ Trp and ^13^C_10_ Kyn in PBMCs were measured by LC-MS (n = 3 technical replicates, representative of n = 1 individual experiment; extracellular proportions of this experiment are found in Fig. S7L-N). Metabolite abundance was normalized to cell numbers. All data are presented as mean ± SEM. Statistical analysis was performed with an unpaired Student’s t- test (A) and with one-way-ANOVA (B-H,J-O). ∗*P* < 0.05; ∗∗*P* < 0.01; ∗∗∗*P* < 0.001, ∗∗∗∗*P* < 0.0001. IDO: Indoleamine 2,3-dioxygenase, Kyn: Kynurenine, QPRT: Quinolinate phosphorybosyltransferase, Trp: Tryptophan.

Next, we confirmed that IFNγ-mediated IDO1 induction depends on JAK/STAT by employing JAK/STAT-inhibitors (Pan-JAK inhibitor: tofacitinib; selective JAK1 inhibitors: upadacitinib and filgotinib; Fig. 6E-H). We subsequently evaluated the metabolic effect of JAK/STAT-activated IDO1 after IFNγ stimulation and found enhanced turnover of Trp to Kyn both in PBMC cell lysates and supernatants, which was reverted upon JAK inhibition (Fig. 6I-K, Fig S7I-K). To further corroborate that increased accumulation of Kyn specifically resulted from the degradation of Trp, we assessed the turnover of the labelled isotope ^13^C_11_ Trp in IFNγ-treated PBMC. Again, IFNγ stimulation led to a significant conversion of ^13^C_11_ Trp into ^13^C_10_ Kyn, which was completely abrogated in response to JAK inhibition (Fig. 6L-N, Fig S6L-N). Altogether, our *in-vitro* data confirm JAK/STAT signalling as a critical driver of mucosal Trp degradation.

### JAK inhibition prevents accumulation of Quin following IFNγ-induced Trp degradation

To investigate the modulatory effect of JAK/STAT on the KP, we sought to further disentangle our observations by detailed profiling of downstream Trp metabolites following IFNγ treatment (Fig. 7A). To this end, human PBMC were subjected to either IFNγ alone or in combination with tofacitinib (JAK inhibitor) or epacadostat (IDO1 inhibitor). Again, IFNγ robustly induced the KP causing a decrease of Trp resulting in heightened Kyn levels (Fig. 7B-G). Augmented abundances of Kyn upon IFNγ treatment led to consecutive increases of anthranilic acid (Anth) and 3-Hydroxyanthranilic acid (3OHAnth) and importantly, to accumulation of Quin (Fig. 7B-G). Conversely, coadministration of tofacitinib or epacadostat effectively abrogated the production of the respective metabolites from Trp, validating that exertion of the KP via IDO1 is JAK/STAT-dependent (Fig. 7B-G). We further measured increased extracellular Kyn, Anth and 3OHAnth secretion in response to IFNγ (Fig. S8B-F). Interestingly, Quin was not detectable in the supernatant of stimulated cells, pointing towards an intracellular accumulation of Quin (Fig. S8B-F).

**Figure 7:**
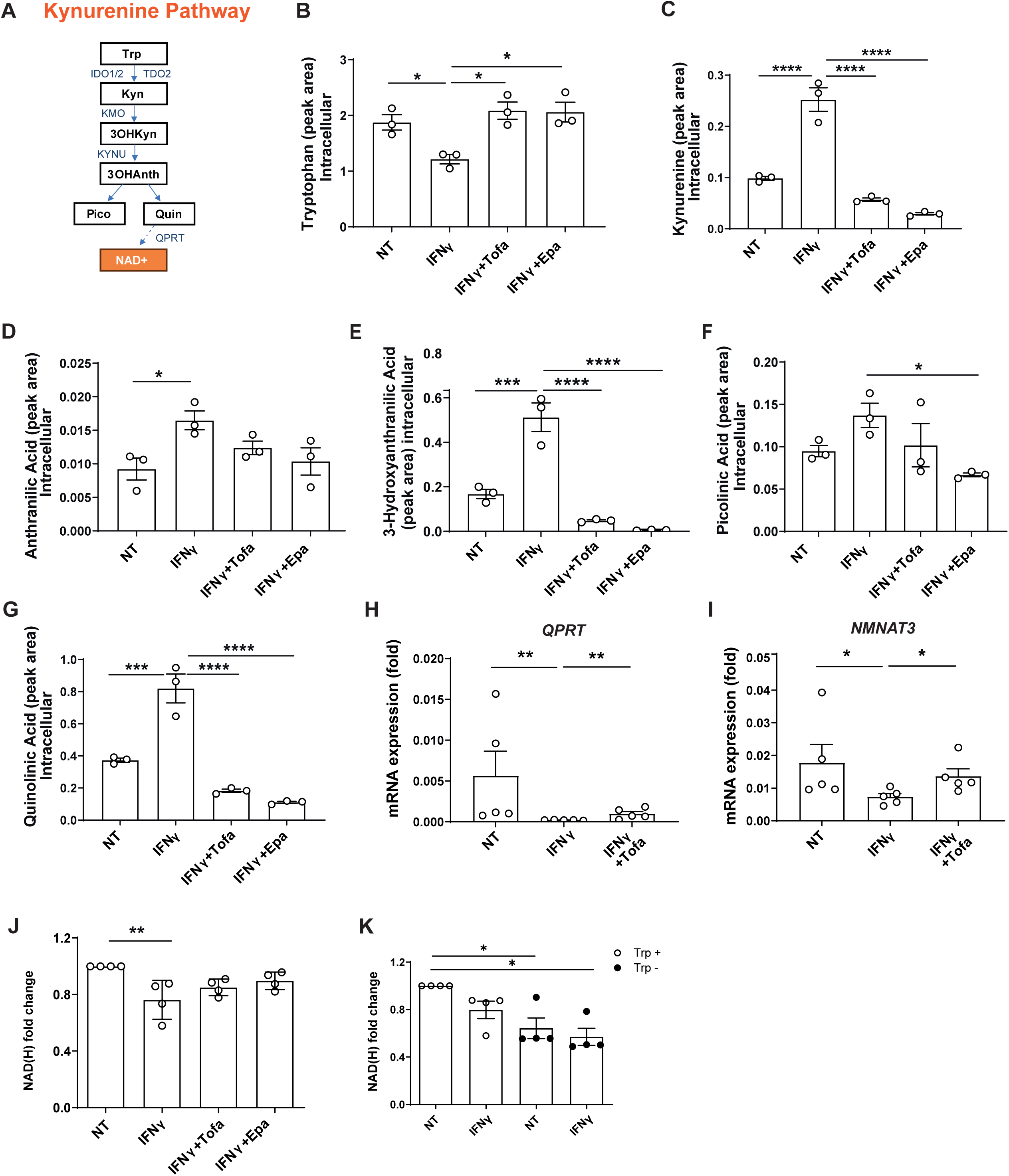
JAK inhibition prevents accumulation of Quin following IFNγ-induced Trp degradation. (A) Overview of the Kynurenine Pathway (KP). Trp: Tryptophan, Kyn: Kynurenine, 3OHKyn: 3- Hydroxy-kynurenine, 3OHAnth: 3-Hydroxyanthranilic acid, Pico: Picolinic acid, Quin: Quinolinic acid, NAD^+^: Nicotinamide adenine dinucleotide. IDO1/2: Indoleamine 2,3- dioxygenase 1/2, TDO: Tryptophan-2,3-dioxygenase, KMO: Kynurenine-3-monooxygenase, KYNU: Kynureninase, QPRT: Quinolinate phosphorybosyltransferase. (B-G) Human PBMC were incubated with IFNγ (10 ng/ml) ± Tofacitinib (Tofa, 1 µM) or Epacadostat (Epa 5 µM) for 24 hours. Depicted metabolites were detected by LC-MS in cell lysis (n = 3 technical replicates, representative of n = 3 individual experiments; metabolite measurements in the respective supernatants are displayed in Fig. S8). Metabolite abundance was normalized to cell numbers. (H,I) Relative gene expression of human PBMC treated with IFNγ (10 ng/ml) ± Tofa (1 µM) for 24 hours was assessed by RT-qPCR (n = 5 technical replicates, representative of n = 2 individual experiments). For gene expression analyses, ß2-Mikroglobulin was used as a housekeeping gene. (J) Human PBMC were incubated with IFNγ (10 ng/ml) ± Tofa (1 µM) or Epa (5 µM) for 24 hours (n = 2 technical replicates, representative of n = 4 individual experiments); (K) human PBMC were incubated with IFNγ (10 ng/ml) in Trp-present (78.4 µg/L) or Trp-deficient (0 µg/L) medium for 24 hours (n = 2 technical replicates, representative of n = 4 individual experiments). For (J) and (K), levels of NAD(H) were measured by NAD/NADH- Glo^TM^ assays (Promega). All data are presented as mean ± SEM. Statistical analysis was performed with one-way-ANOVA. ∗*P* < 0.05; ∗∗*P* < 0.01; ∗∗∗*P* < 0.001; ∗∗∗∗*P* < 0.0001. IDO1/2: Indoleamine 2,3-dioxygenase 1/2, KMO: Kynurenine-3-monooxygenase, Kyn: Kynurenine, KYNU: Kynureninase, NAD^+^: Nicotinamide adenine dinucleotide, NMNAT3: Nicotinamide Nucleotide Adenylyltransferase 3, Pico: Picolinic acid, QPRT: Quinolinate phosphorybosyltransferase, Quin: Quinolinic acid, Trp: Tryptophan, TDO: Tryptophan-2,3- dioxygenase, 3OHKyn: 3-Hydroxy-kynurenine, 3OHAnth: 3-OH-Anthranilic acid.

We next questioned whether Quin accumulation following IFNγ stimulation in PBMC lysates was caused by a blockade of the KP at the level of QPRT. In line with our previous findings, gene expression of *QPRT* was thoroughly suppressed upon IFNγ stimulation, whereas tofacitinib cotreatment lead to a recovery of *QPRT* expression (Fig. 7H). Of note, treatment with LPS and IFNγ in comparison to other cytokines led to the strongest suppression of *QPRT* expression (Fig. 7H, S8G,H). Altogether, our findings suggest a so far undescribed role of IFNγ in inducing not only the KP via IDO1, but also a blockade in the conversion of Quin to NAD^+^.

Based on these findings, we hypothesized that reduced *QPRT* gene expression would result in decreased conversion of Quin into NAD^+^. Indeed, we found NAD(H) production from Trp in human PBMC to be decreased upon stimulation with IFNγ (Fig. 7J,K). An even stronger depletion of cellular NAD(H) abundances was observed in the context of Trp-deficient medium (Fig. 7K). We concluded that conversion of Quin to NAMN by QPRT is crucial for NAD^+^ synthesis. In line with this hypothesis, nicotinamide nucleotide adenylyltransferase 3 (*NMNAT3*), the enzyme processing the Quin-derivative NAMN, was suppressed upon IFNγ administration (Fig. 7I), which may suggest a lack of available substrates.

Concluding, we found that IFNγ-driven Trp degradation along the KP resulted in accumulation of Quin due to decreased *QPRT* expression, leading to reduced cellular bioavailability of NAD^+^.

### QPRT coordinates cellular inflammatory responses

After resolving the impact of JAK/STAT-driven Trp degradation in the myeloid compartment, we set out to understand the consequences of downregulated *QPRT* on cellular homeostasis. As QPRT was predominantly expressed in fibroblasts, where it was found to be strongly downregulated within the inflamed CD mucosa (Fig. 5G), we used a fibroblast (CCD18) cell culture model and silenced *QPRT* with siRNA. Knockdown of *QPRT* significantly increased gene expression of pro-inflammatory cytokines upon stimulation with IFNγ and/or LPS (Fig. 8A-F). We next assessed potential direct cytotoxic effects of external Quin stimulation on cytokine expression and cell viability, as Quin might be released from cells with low *QPRT* expression, exposing neighboring intestinal lineages to high levels of Quin. Even at high concentrations, Quin neither amplified pro-inflammatory cytokine responses nor significantly reduced cellular viability of PBMC (Fig. S9A-D), epithelial cells (Fig. S9E-H) or fibroblasts (Fig. S9I-K), arguing against direct effects of extracellular Quin on promoting inflammation. To mechanistically assess whether knockdown of *QPRT* drives pro-inflammatory cytokine expression via impaired *de-novo* NAD^+^ synthesis, we assessed cytokine responses and NAD^+^ levels following treatment with IFNγ and/or LPS in combination with tofacitinib or epacadostat to block KP activation. In addition, we conducted co-treatment with the NAD^+^ precursor nicotinamide riboside (NR) ^37^. IFNγ-and/or LPS-induced increases of pro-inflammatory cytokines in *QPRT*-silenced cells were not ameliorated by epacadostat, further arguing against a pathological accumulation of intracellular quinolinic acid as driver of inflammation. In contrast they were, however, rescued by tofacitinib and NR supplementation (Fig. 8D-F). At the same time, NAD(H) levels were significantly reduced upon IFNγ and/or LPS stimulation, which was further aggravated by concomitant knockdown of *QPRT* (Fig. 8G). Only supplementation of NR could restore NAD(H) levels in cells treated with IFNγ and/or LPS. Co-treatment with tofacitinib or epacadostat did not significantly elevate NAD(H) abundances, presumably due to reduced flow of Trp into the KP, thereby reducing *de-novo* NAD^+^ synthesis rates (Fig. 8G). Altogether, these data corroborate that reduction of metabolic flow from Trp via QPRT drives a pro-inflammatory cellular state in immuno-competent cell types of the intestinal mucosa. This can largely be overcome by substitution of NAD^+^ precursors or by promoting anti-inflammatory signalling and thereby lowering cellular requirement of NAD^+^.

**Figure 8:**
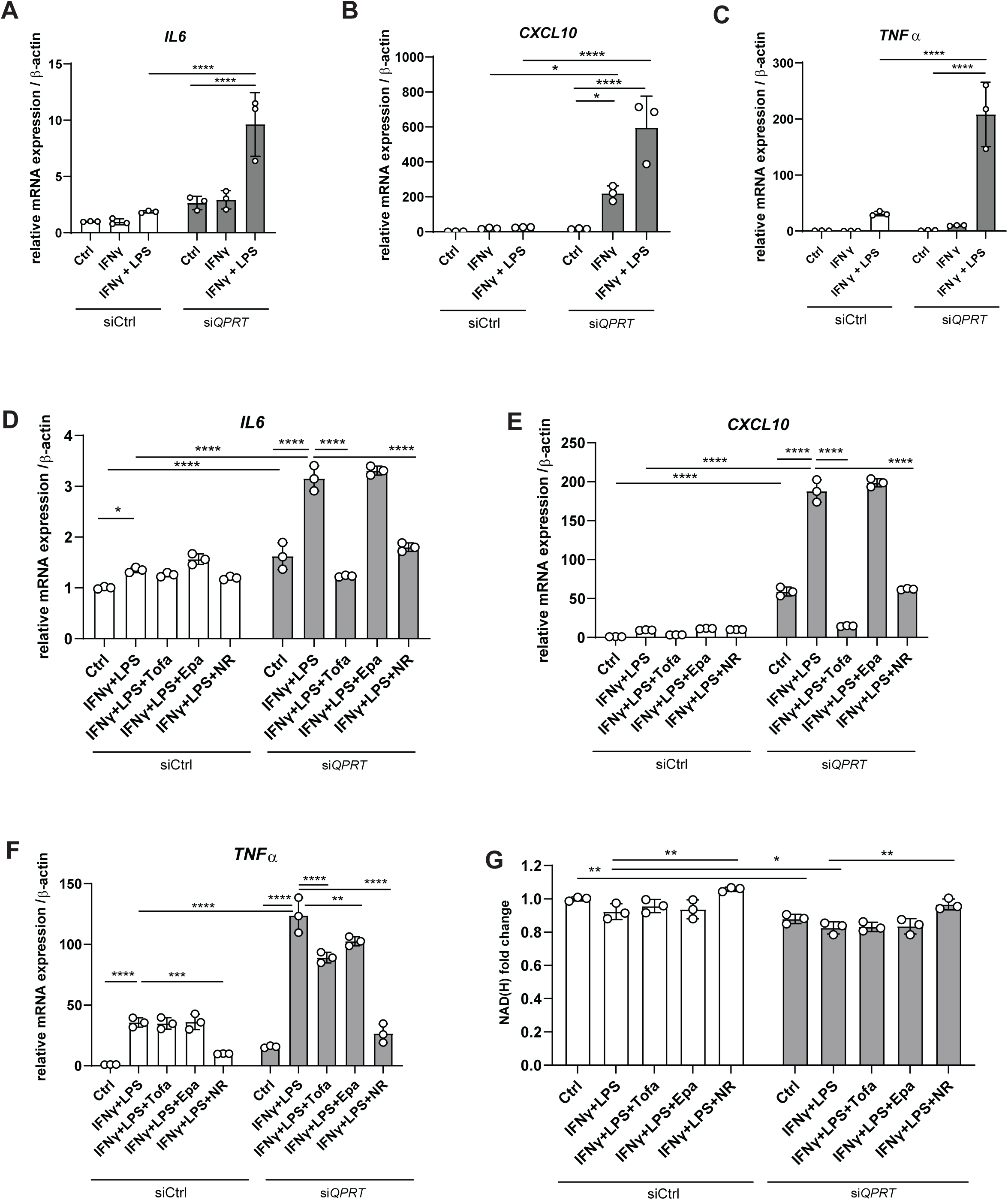
QPRT coordinates cellular inflammatory responses. Human fibroblasts were stimulated (A-C) with IFNy (10 ng/ml for 24 hours) and in combination with IFNy and LPS (1 µg/ml for 2 hours), whereas in (D-G), cells where additionally treated with Tofacitinib (Tofa, 1 µM, 24 hours), Epacadostat (Epa, 5 µM, 24 hours) or Nicotinamide Riboside (NR, 10 mM, 24 hours). (G) NAD(H) levels were measured via NAD Glo Assay. To knockdown *QPRT*, cells were transfected with siRNA against *QPRT*. Gene expression was assessed by RT-qPCR, ß-Actin served as a housekeeper. n = 3 technical replicates, representative of n = 3 individual experiments. Data are presented as mean ± SEM, statistical analysis with one-way ANOVA. ∗P < 0.05; ∗∗P < 0.01; ∗∗∗P < 0.001; ∗∗∗∗P < 0.0001. QPRT: Quinolinate phosphorybosyltransferase.

## Discussion

Increased degradation of tryptophan (Trp) along the kynurenine pathway (KP) has been described as a metabolic principle in several immune-mediated diseases to overcome cellular energy deprivation. Here, we investigated the directionality of the KP and show that in the context of IBD, where excessive KP activation is observed, catabolism of Trp is skewed by an inefficient NAD^+^ *de-novo* synthesis.

Starting with a clinical observation, we found that IBD patients undergoing successful targeted therapy exhibited a steady increase in their serum Trp levels over 52 weeks. We reasoned that Trp restoration is a result of decreased Trp catabolism along the KP. Given the augmented demand for NAD^+^ in inflammatory settings and the presence of niacin deficiency in IBD ^38–40^, it is reasonable to assume that Trp catabolism is increased in active IBD to amplify *de-novo* NAD^+^ synthesis. However, despite strong activation of the KP in IBD, we observe heightened levels of the NAD^+^ precursor quinolinic acid (Quin) correlating with Kyn:Trp ratios in the serum of active IBD patients, suggesting reduced Quin-to-NAD^+^ conversion. Importantly, increases in Quin have previously been observed in the context of IBD, rheumatoid arthritis and other inflammatory as well as neuro-degenerative diseases ^20,41–43^. To acknowledge that sex-specific differences in Trp metabolism exist ^22^, we included sex assigned at birth as a covariate in our models. However, an individual assessment of sexes may provide additional important information.

Examining the mucosal and blood transcriptome of IBD patients, we found that a major proportion of Trp catabolism occurs in the inflamed mucosa. Notably, the strong mucosal upregulation of the KP appeared to encounter a roadblock at Quin, where quinolinate phosphorybosyltransferase (QPRT) gene and protein expression was downregulated in correlation with augmented endoscopic and histologic disease activity. To rule out that serum Quin accumulation might originate from other organs, we conducted targeted metabolomics of Trp metabolites in several immunoactive anatomical compartments in the context of DSS colitis and validated that i) Trp degradation along the KP occurs almost exclusively at the site of active inflammation, where ii) it is characterized by a local increase in Quin and reduction of NAD(H). Although the DSS model is an acknowledged preclinical IBD model, one limitation of our study is that targeted metabolomics of Trp metabolism have not yet been conducted on human IBD biopsies to validate the compartment specific degradation of tryptophan in IBD. To identify the upstream activator of the KP in IBD, we conducted integrated transcriptome and metabolome analysis and identified the janus kinase / signal transducers and activators of transcription (JAK/STAT) pathway as a major regulator of Trp degradation in a disease-dependent manner ^31,33,44^. We demonstrated that primarily IFNγ, but also other pro-inflammatory cytokines signalling via JAK/STAT, including IL-23 and TNFα, induced *IDO1* in human PBMC. Paradoxically, these cytokines impaired expression of *QPRT,* in line with previous work in macrophages demonstrating downregulation of *de-novo* NAD^+^ synthesis by LPS, which drives a pro-inflammatory state due to insufficient NAD^+^ abundance ^21^. Importantly, several cytokines capable of JAK/STAT activation have been described to impair mitochondrial function and to induce a metabolic switch towards the NAD^+^ salvage pathway, as an attempt to replenish NAD^+^ levels and facilitate redox balance ^45,46^. JAK inhibitors (JAKi) are established therapies in several chronic inflammatory disorders, including IBD, and it has been shown that

JAKi restore cellular ATP levels and mitochondrial function ^47–54^. Based on our cellular models, JAKi might exert a combined beneficial effect by i) lowering the overall cellular energy demand, ii) preventing the build-up of Quin and iii) by ameliorating the metabolic constraint of reduced *QPRT* expression. Notably, this hypothesis is supported in our finding that UC patients with high baseline *IDO1* and low baseline *QPRT* expression were more sensitive to JAKi and therefore had a higher chance to respond to JAKi therapy (Fig. 4L). As it has previously been described that the microbiome mediates intestinal homeostasis upon mitochondrial pertubations ^55^, a limitation of our study is that the impact of the intestinal microbiome on inflammation-associated metabolism has not yet been considered.

Having established the critical role of QPRT in intestinal inflammation, the question remains whether this phenomenon depends on direct cytotoxic effects of Quin. Accumulation of Quin has been ascribed pro-inflammatory properties in the context of rheumatoid arthritis and is an established neurotoxin ^41,56^. Quin is furthermore associated with increased oxidative stress, ATP exhaustion and disturbed mitochondrial function as well as with diminished lifespan of *C. elegans* ^57,58^. However, we were not able to identify a pro-inflammatory effect following exogenous Quin stimulation in epithelial cells, PBMC or fibroblasts. In contrast, when knocking down *QPRT* with siRNA in fibroblasts, which we identified in scSeq analysis as the cell type most strongly affected by inflammation-induced *QPRT* downregulation, we observed an increase of pro-inflammatory cytokines that was further augmented following stimulation with IFNγ/LPS. Supplementation with NR could to great extent rescue the pro-inflammatory phenotype, suggesting that both intracellular Quin flooding, but predominantly NAD^+^ exhaustion promoted the aggravated inflammatory response. This was in line with our finding that treatment with the IDO1 inhibitor epacadostat, despite its inhibitory effect on Quin accumulation, could not rescue the hyperinflammatory state following IFNγ and/or LPS stimulation, as it also causes NAD^+^ depletion. Hence, our data highlight the critical function of cell type-specific conversion of Quin to NAD^+^. Further research is required to examine intercellular exchanges of KP metabolites. Given the knowledge that NAD^+^ repletion can directly improve mitochondrial function and cellular fitness ^59,60^, it might be assumed that the insufficient provision of NAD^+^ via QPRT might impact stem cell function and wound repair in intestinal inflammation.

Altogether, our data underscore the critical role of fueling cellular energy levels via KP-mediated *de-novo* NAD^+^ synthesis. Therapeutic strategies should therefore aim to restore cellular NAD^+^ levels directly at the site of NAD^+^ shortage (e.g., inflamed mucosa), such as by ileocolonic release of NAD^+^ precursors (NCT05258474) or targeted restoration of QPRT function.

## Data Availability

All data produced in the present study are available upon reasonable request to the authors

## Acknowledgements

We would like to appreciate the excellent work of our technical assistants Christina Nimke, Janina Ohrndorf, Meike Hansen, Ronja Möhring, Sophie Reiher, Sabine Kock, Tanja Klostermeier, Stefanie Rentzow and Dorina Ölsner. We would further like to extend our sincere gratitude to the patients who participated in this study, without whom our research would not be possible. Lastly, we would like to acknowledge the Huck Institutes’ Metabolomics Core Facility (RRID:SCR_023864) for use of the OE 240 LC-MS and Sergei Koshkin for helpful discussions on sample preparation and analysis.

## Disclosures

The authors have no conflict of interest.

## Author Contributions

K.A., P.R., L.W., D.H., N.K., A.I.A. designed the study.

N.K., A.I.A., L.W., D.H., Q.W., M.O., J.T., G.C., V.V., P.M., F.R., E.K., F.T., L.K.S., P.P., N.P.,

S.W. performed experiments and analysed the data.

P.R., K.A., M.R.M., E.L., B.V., F.F., C.K. planned the project and supervised the experiments. L.W., D.H., N.K., K.A., P.R. wrote the initial manuscript.

K.A., M.R.M., P.R., S.Schr. edited the manuscript

## Abbreviations

3OHAnth: 3-Hydroxyanthranilic acid
3OHKyn: 3-Hydroxy-kynurenine
5OHTrp: 5-Hydroxy-tryptophan
AADAT: adenosine deaminase acting on tRNA
Anth: Anthranilic acid
Epa: Epacadostat
Filgo: Filgotinib
HIAA: 5-Hydroxy indoleacetic acid
IDO1/2: Indoleamine 2,3-dioxygenase ½
IFNγ: Interferon gamma
IL: Interleukin
IPA: Indole-3-propionic acid
JAK/STAT: Janus kinase / signal transducers and activators of transcription
KP: Kynurenine pathway
KMO: Kynurenine-3-monooxygenase
Kyn: Kynurenine
Kyna: Kynurenic acid
KYNU: Kynureninase
LPS: Lipopolysaccharides
NA: Nicotinic acid
NAD^+^: Nicotinamide adenine dinucleotide
NAM: Nicotinamide
NAMN: Nicotinic acid mononucleotide
NAPRT: Nicotinic acid phosphoribosyltransferase
Neopt: Neopterin
NMNAT3: Nicotinamide nucleotide adenylyltransferase 3
NR: Nicotinamide riboside
Pico: Picolinic acid
QPRT: Quinolinate phosphorybosyltransferase
Quin: Quinolinic acid
Sero: Serotonin
TDO: Tryptophan-2,3-dioxygenase
TNFα: Tumor necrosis factor alpha
Tofa: Tofacitinib
Trp: Tryptophan
Upa: Upadacitinib
Xanth: Xanthurenic acid

## Figure Legends

**Figure S1:**
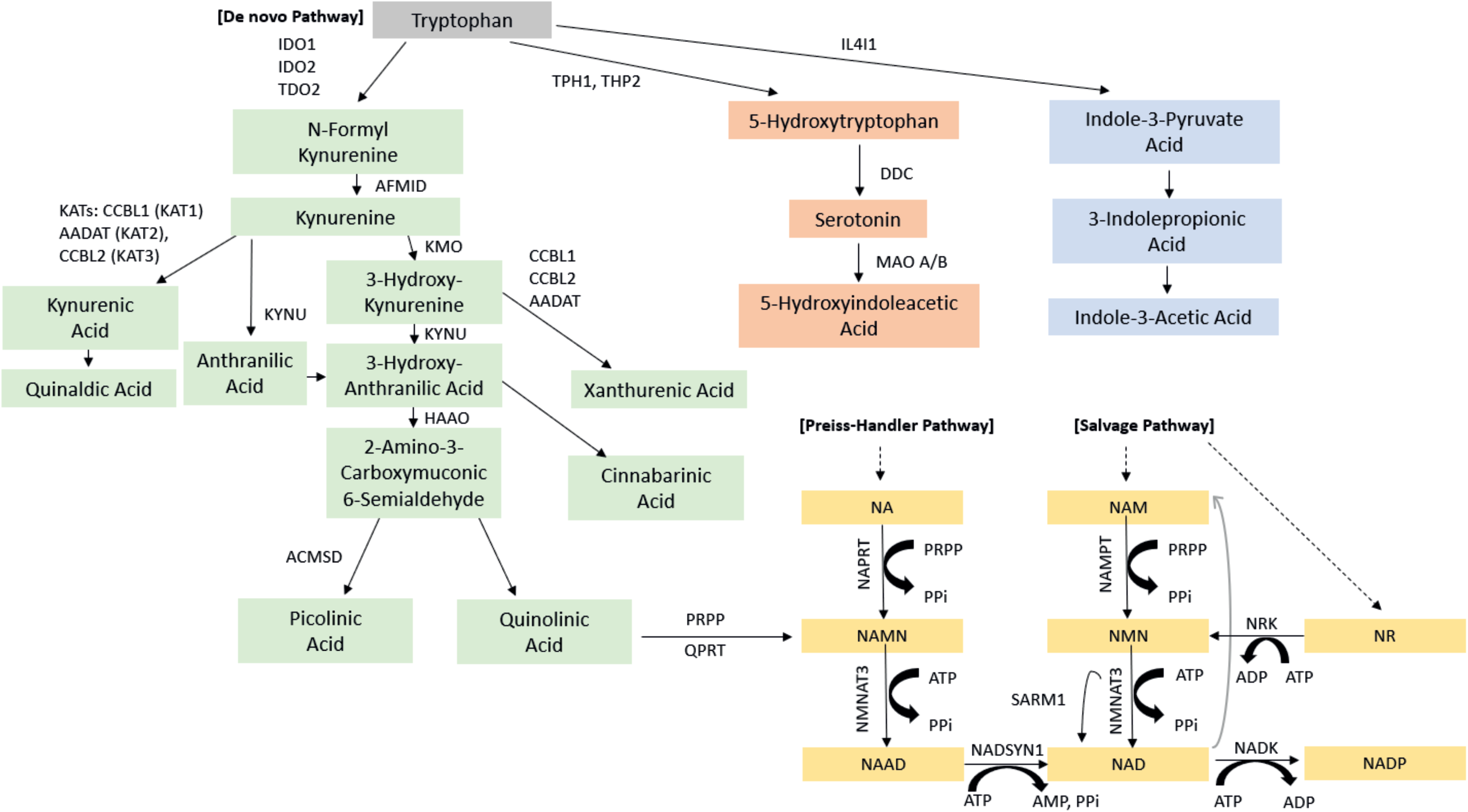
Overview of Trp degrading pathways and NAD^+^ synthesis from *de-novo* synthesis, Preiss-Handler pathway and salvage pathway. AADAT: Kynurenine/alpha-aminoadipate aminotransferase, ACMSD: Aminocarboxymuconate semialdehyde decarboxylase, ADP: Adenosine diphosphate, AFMID: Arylformamidase, ATP: Adenosine triphosphate, CCBL: Cysteine conjugate beta-lyase, HAAO: 3-Hydroxyanthranilate 3,4-dioxygenase, DDC: Dopa decarboxylase, IDO: Indoleamine 2,3-dioxygenase, IFNγ: Interferon gamma, IL4I1: Interleukin-4-induced-1, IL-23: Interleukin 23, KAT: Kynurenine aminotransferase, KMO: Kynurenine-3-monooxygenase, KYNU: Kynureninase, NA: Nicotinic acid, NAAD: Nicotinic acid adenine dinucleotide, NAD: Nicotinamide adenine dinucleotide, NADP: Nicotinamide adenine dinucleotide phosphate, NADK: Nicotinamide adenine dinucleotide kinase, NADSYN1: Nicotinamide adenine dinucleotide synthetase 1, NAM: Nicotinamide, NAMN: Nicotinic acid mononucleotide, NAMPT: Nicotinamide phosphoribosyltransferase, NAPRT: Nicotinate phosphoribosyltransferase, NNMT: Nicotinamide N-methyltransferase, NMN: Nicotinamide mononucleotide, NMNAT3: Nicotinamide nucleotide adenylyltransferase 3, NR: Nicotinamide riboside, NRK: Nicotinamide riboside kinase, PPi: Pyrophosphate, PRPP: Phosphoribosyl pyrophosphate, QPRT: Quinolinate phosphorybosyltransferase, SARM1: sterile alpha and TIR motif containing 1, TDO2: Tryptophan-2,3-dioxygenase 2, TPH: Tryptophan hydroxylase, TNFα: Tumor necrosis factor alpha.

**Figure S2:**
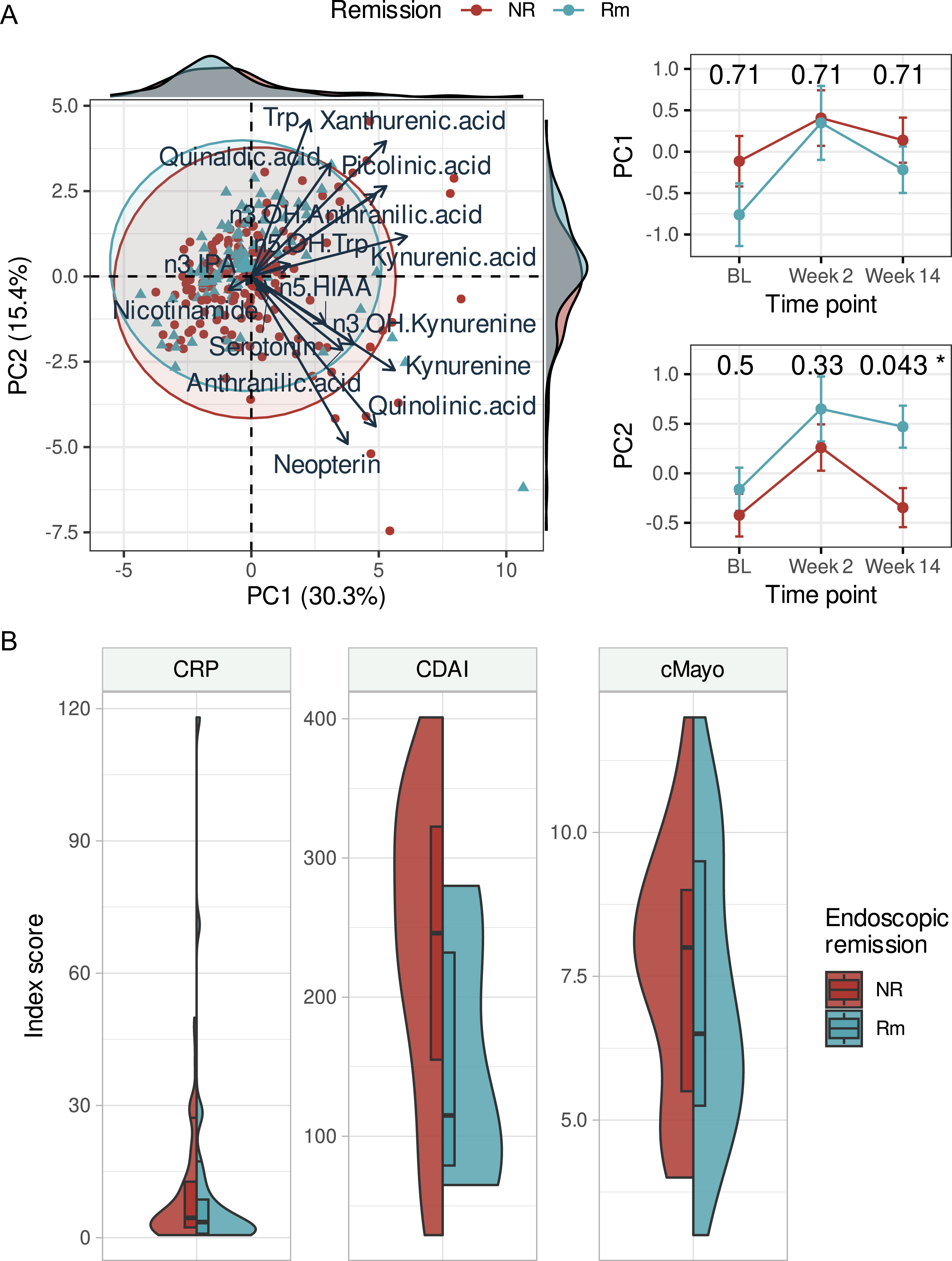
Principal component (PC) analysis of Trp-derivatives and baseline disease activity. (A) We observed changes over time in PC2, but not PC1, for patients in endoscopic remission compared to those who did not enter remission. (B) No statistically significant changes in baseline activity were observed between individuals who entered endoscopic remission at week 14, but there was a clear trend for heightened disease activity among individuals who would enter remission. Endoscopic remission was determined at week 14 based on available endoscopic Mayo (< 2) or SES-CD (< 3). In total, 34 patients entered remission by week 14 and 57 did not. Rm: Remission, NR: Non-remission.

**Figure S3:**
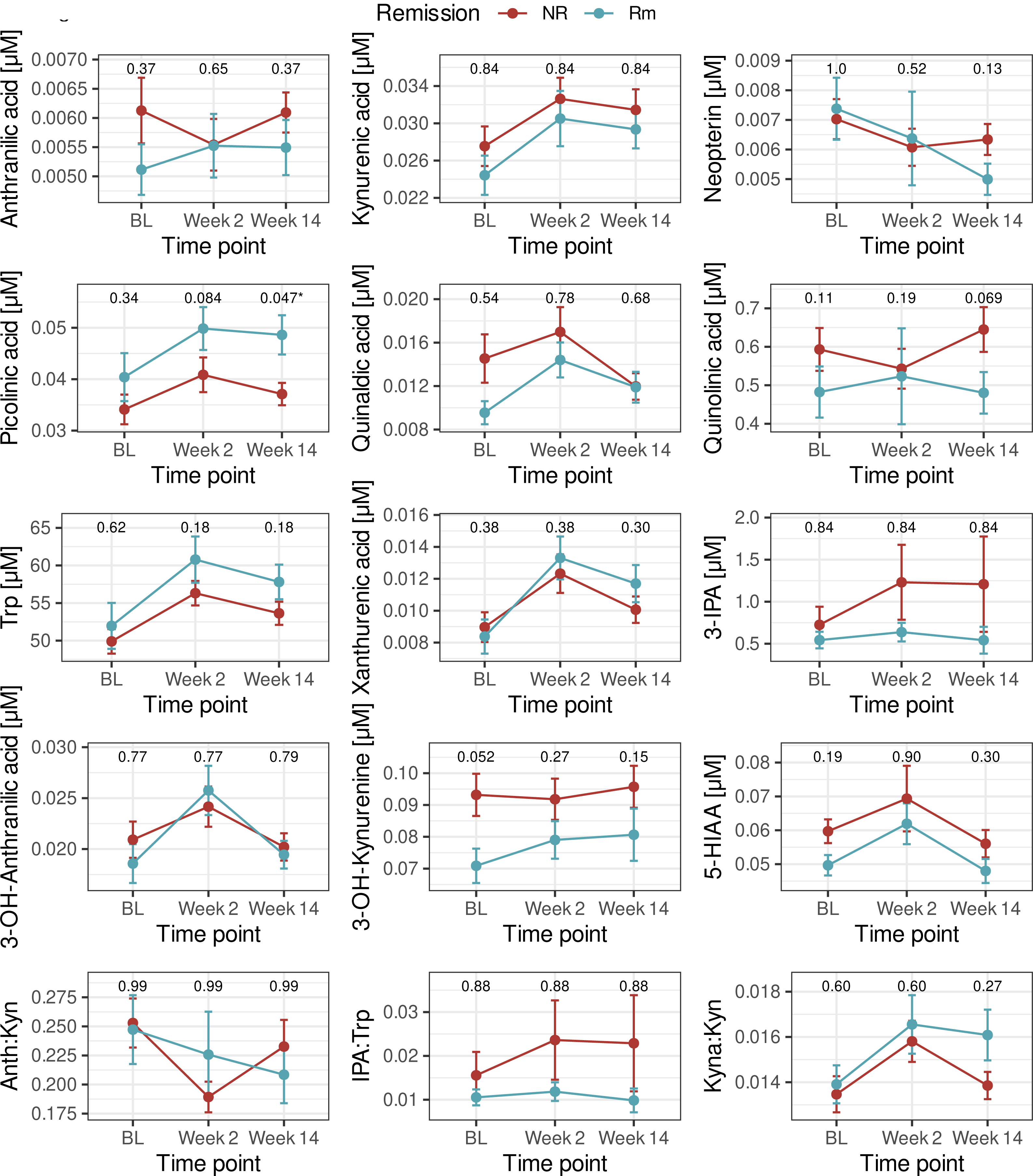
Line plots: Trajectories for the remaining metabolites and metabolite ratios that exhibited significant relationships with at least one disease activity metric. Remission was determined at week 14 (endoscopic Mayo < 2 or SES-CD < 3). Total patients per category were 34 remission (Rm) and 57 non-remission (NR). Neopt: Neopterin, Quin: Quinolinic acid, HIAA: 5-Hydroxy indoleacetic acid, 3OHKyn: 3-Hydroxy-kynurenine, 5OHTrp: 5-Hydroxy-tryptophan, Sero: Serotonin, Kyn: Kynurenine, Nicotin: Nicotinamide, Anth: Anthranilic acid, 3OHAnth: 3-OH-Anthranilic acid, IPA: Indole-3-propionic acid, Kyna: Kynurenic acid, Pico: Picolinic acid, Trp: Tryptophan, Xanth: Xanthurenic acid, Quinald: Quinaldehyde.

**Figure S4:**
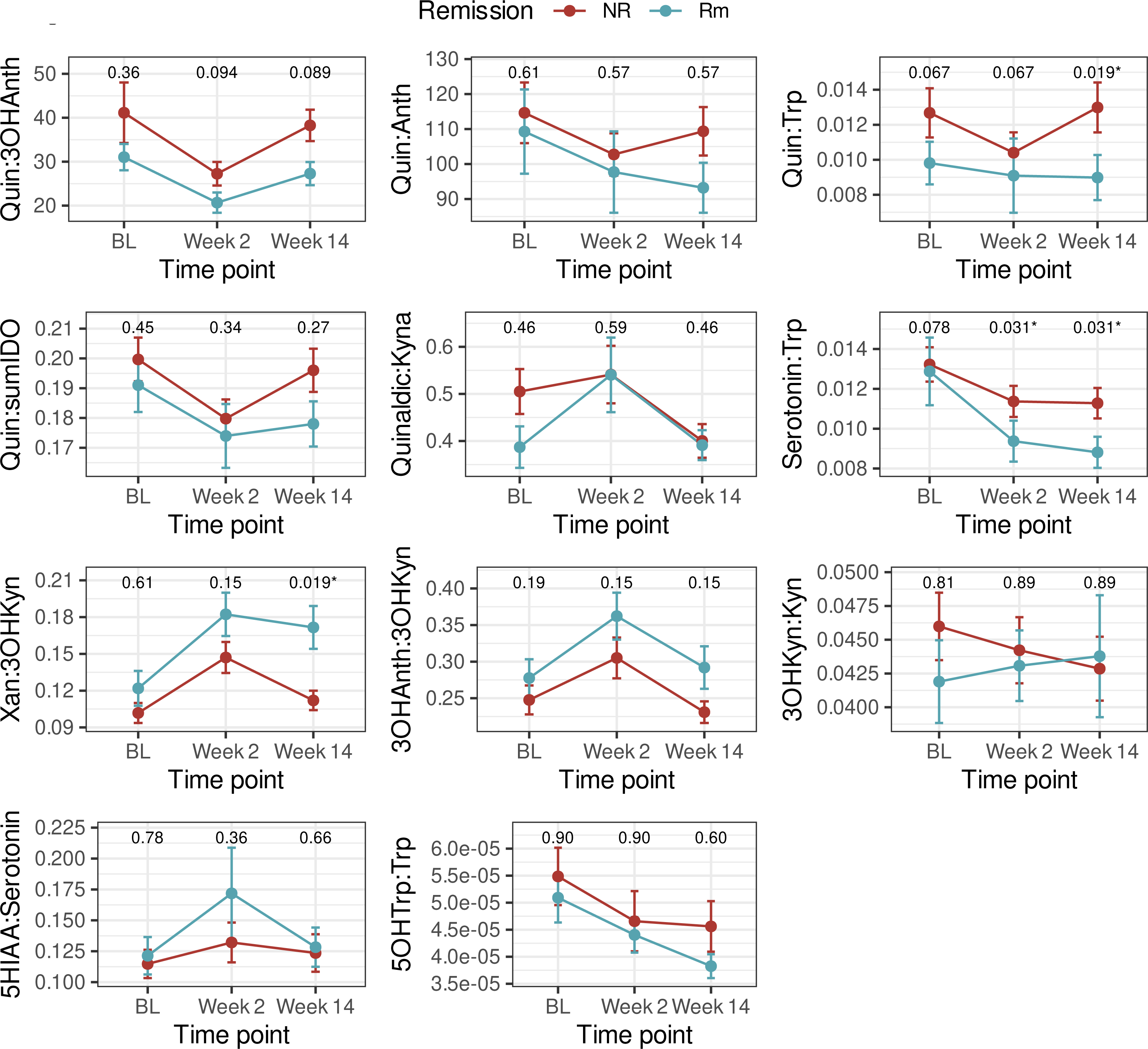
Line plots: Trajectories for the remaining metabolites and metabolite ratios that exhibited significant relationships with at least one disease activity metric. Remission was determined at week 14 (endoscopic Mayo < 2 or SES-CD < 3). Total patients per category were 34 remission (Rm) and 57 non-remission (NR). Neopt: Neopterin, Quin: Quinolinic acid, HIAA: 5-Hydroxy indoleacetic acid, 3OHKyn: 3-Hydroxy-kynurenine, 5OHTrp: 5-Hydroxy-tryptophan, Sero: Serotonin, Kyn: Kynurenine, Nicotin: Nicotinamide, Anth: Anthranilic acid, 3OHAnth: 3-OH-Anthranilic acid, IPA: Indole-3-propionic acid, Kyna: Kynurenic acid, Pico: Picolinic acid, Trp: Tryptophan, Xanth: Xanthurenic acid, Quinald: Quinaldehyde.

**Figure S5:**
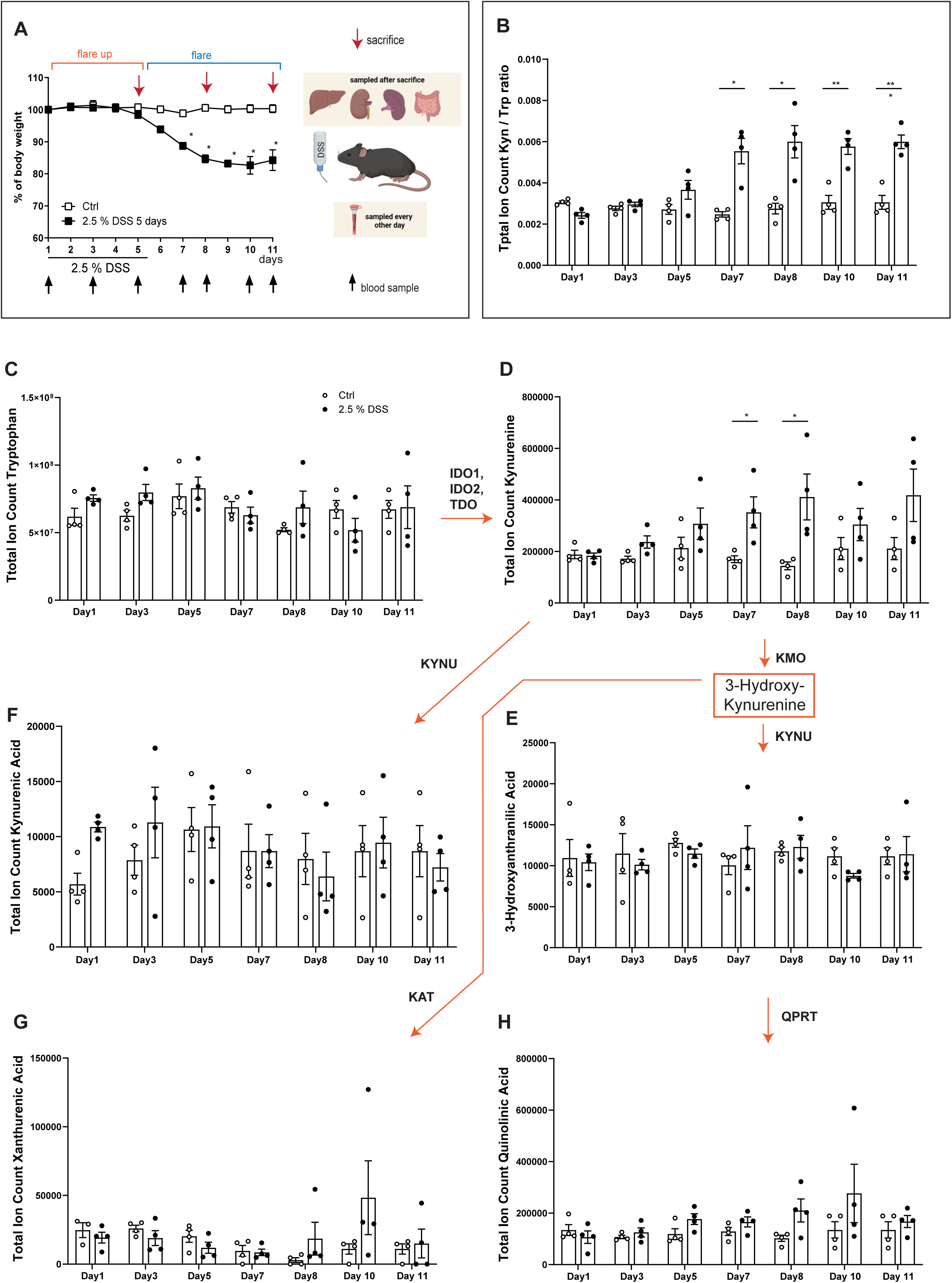
Mucosal Trp degradation is reflected in serum during DSS colitis. (A) Experimental set-up: 8-12 weeks old, male C57BL/6J mice (n = 8) were supplied with drinking water (white, control) or 2.5% DSS for 5 days (black, normal drinking water was supplied from day 6 on). Blood was collected every over day (indicated by black arrows). (B) Kyn/Trp ratio of the metabolites measured in (C) and (D). (B-H) Total ion counts of Trp metabolites were measured by LC-MS in murine serum sampled every other day during DSS colitis. Groups: <u>White</u> = control groups (n = 4); <u>black</u>: DSS-treated animals (n = 4). Each dot represents the measurements of one individual mouse. All data are presented as mean ± SEM. Statistical analysis was performed with Mann-Whitney-U test. ∗*P* < 0.05; ∗∗*P* < 0.01; ∗∗∗*P* < 0.001; ∗∗∗∗*P* < 0.0001. HAAO: 3-Hydroxyanthranilate 3,4-dioxygenase, IDO1: Indoleamine 2,3-dioxygenase 1, KAT: Kynurenine aminotransferases, KMO: Kynurenine-3- monooxygenase, Kyn: Kynurenine, KYNU: Kynureninase, QPRT: Quinolinate phosphorybosyltransferase, TDO: Tryptophan-2,3-dioxygenase, Trp: Tryptophan.

**Figure S6:**
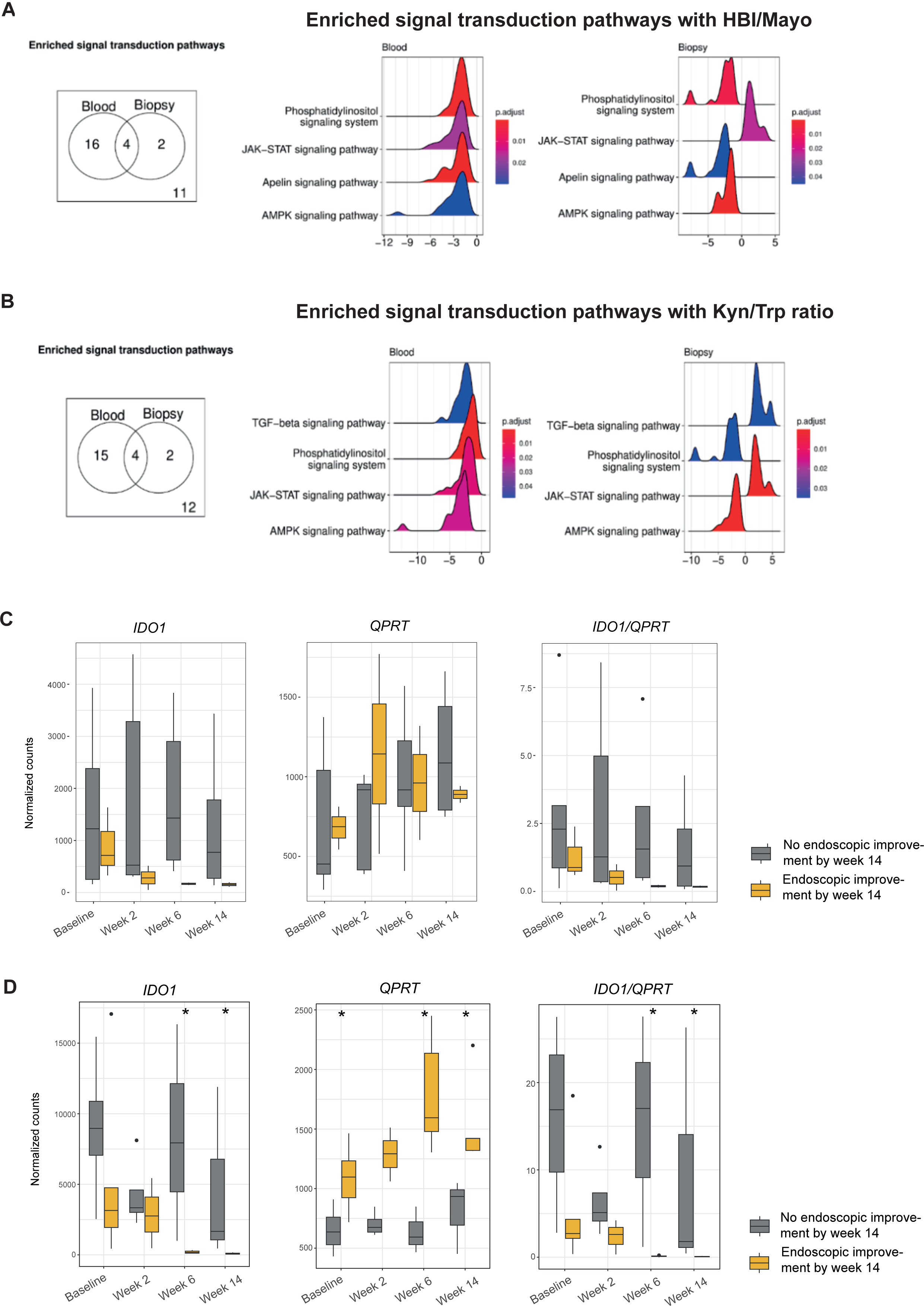
JAK/STAT drives mucosal Trp degradation in IBD. (A,B) Whole blood and biopsies were obtained within two longitudinal IBD intervention cohorts sampled in northern Germany (n = 62 IBD, n = 32 UC, n = 30 CD) over a period of 14 weeks at the indicated timepoints. Patients were treatment-naïve for biologic treatment and were introduced to either anti-TNFα (n = 22), anti-α4β7-Integrin (n = 21), anti-IL6-trans (n = 16) or anti-IL6-R (n = 3). Transcriptomics were conducted on whole blood and biopsies, and serum samples were used for metabolomics. Differential gene expression analysis was performed using linear mixed models to associate changes in gene expression in biopsies and blood with changes in (A) disease activity (HBI/Mayo) and (B) serum Kyn/Trp ratios. Afterwards, we used gene set enrichment (GSEA) and hypergeometric tests on the signal transduction pathways annotated by KEGG. All p-values in this figure were adjusted with the Benjamini-Hochberger correction. (C,D) The normalized expression of *IDO1*, *QPRT* and *IDO1*/*QPRT* ratio in mucosal biopsies of UC patients was assessed for different biological treatments. Endoscopic improvement was defined as a Mayo endoscopic subscore ≤1, as described before ^62^. (C) UC patients were treated with infliximab within a longitudinal IBD cohort study. A total of 28 longitudinal samples were included in the analysis (baseline = 8; week 2 = 7; week 6 = 7; week 14 = 6). (D) UC patients were treated with vedolizumab within a longitudinal IBD cohort study. A total of 32 longitudinal samples were included in the analysis (baseline = 9; week 2 = 7; week 6 = 8; week 14 = 8). For (C,D), the Wilcoxon test was used to determine significant difference between responders and non-responders at each time point. *p-value ≤ 0.05. IDO1: Indoleamine 2,3-dioxygenase 1, Kyn: Kynurenine, QPRT: Quinolinate phosphorybosyltransferase, Trp: Tryptophan.

**Figure S7:**
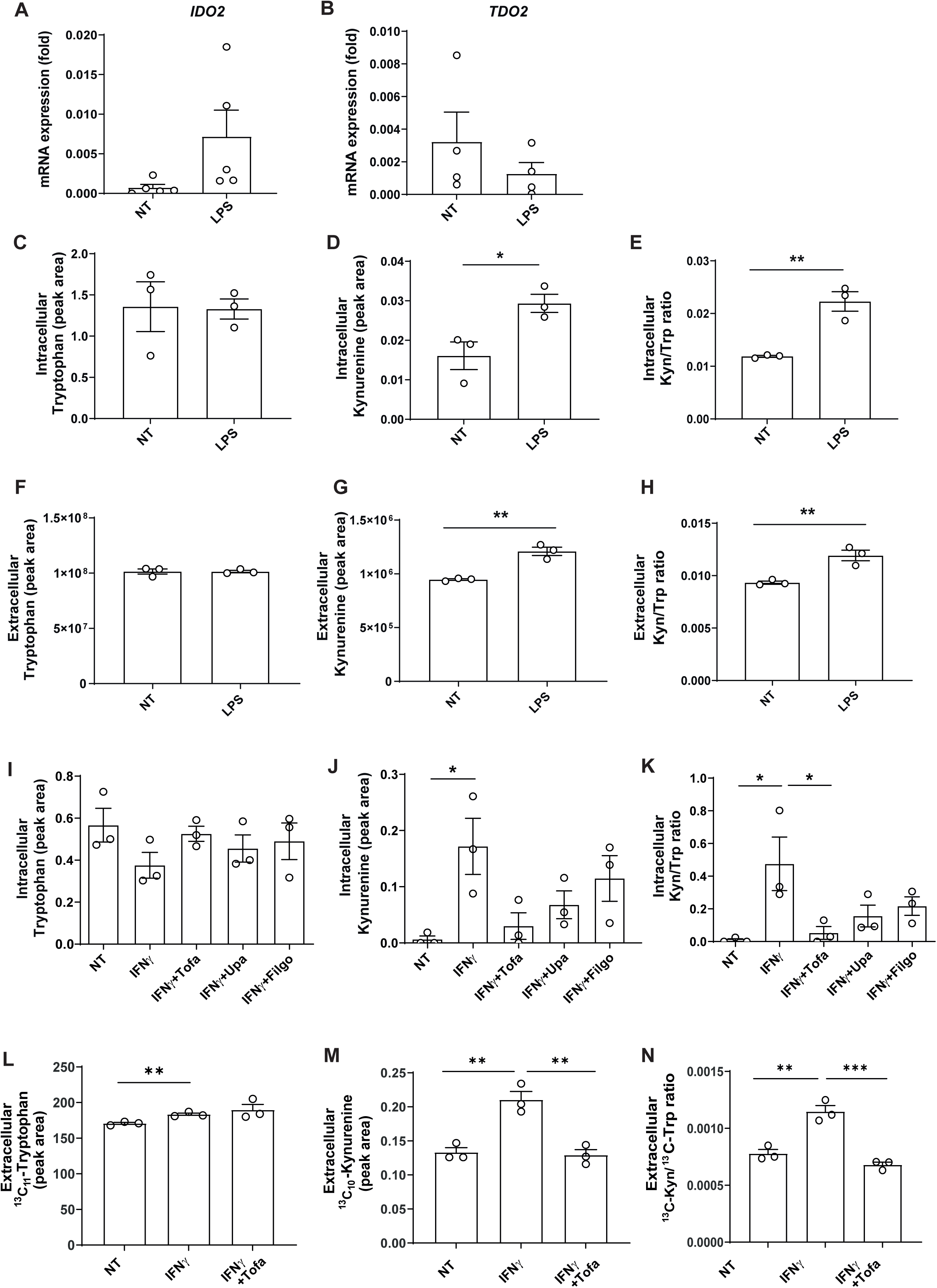
IFNγ-driven Trp conversion via IDO1 is restored by JAK inhibition. Human PBMC were treated with LPS (100 ng/ml) for 24 hours. (A,B) Relative gene expression was assessed by RT-qPCR (n = 2 technical replicates, representative of n = 5 (*IDO2*) or n=4 (*TDO2*) individual experiments; part of the gene expression analysis found in Fig. 6A). ß2- Mikroglobulin was used as housekeeping gene. (C-E) Intra- and (F-H) extracellular Trp and Kyn metabolites levels were assessed by LC-MS (n = 3 technical replicates, representative of n = 3 individual experiments). (I-K) Human PBMC were incubated with IFNγ (10 ng/ml) ±1 µM tofacitinib (Tofa), 0.1 µM upadacitinib (Upa) or 10 µM filgotinib (Filgo) for 24 hours. Intracellular Trp and Kyn levels were measured by LC-MS (n = 3 technical replicates, representative of n = 3 individual experiments; extracellular proportions of this experiment are found in Fig. 6I-K). (L-N) Human PBMC were incubated with IFNγ (10 ng/ml) ± Tofa (1 µM) for 24 hours. Extracellular levels of ^13^C_11_ Trp and ^13^C_10_ Kyn in PBMCs were measured by LC-MS (n = 3 technical replicates, representative of n = 1 individual experiment; intracellular proportions of this experiment are found in Fig. 6L-N). Metabolite abundance was normalized to cell numbers. All data are presented as mean ± SEM. Statistical analysis was performed with an unpaired Student’s t-test for (A-H) or one-way-ANOVA (I-N). ∗*P* < 0.05; ∗*P* < 0.05; ∗∗*P* < 0.01; ∗∗∗*P* < 0.001. Kyn: Kynurenine, Trp: Tryptophan. IDO: Indoleamine 2,3-dioxygenase, QPRT: Quinolinate phosphorybosyltransferase, TDO: Tryptophan-2,3-dioxygenase.

**Figure S8:**
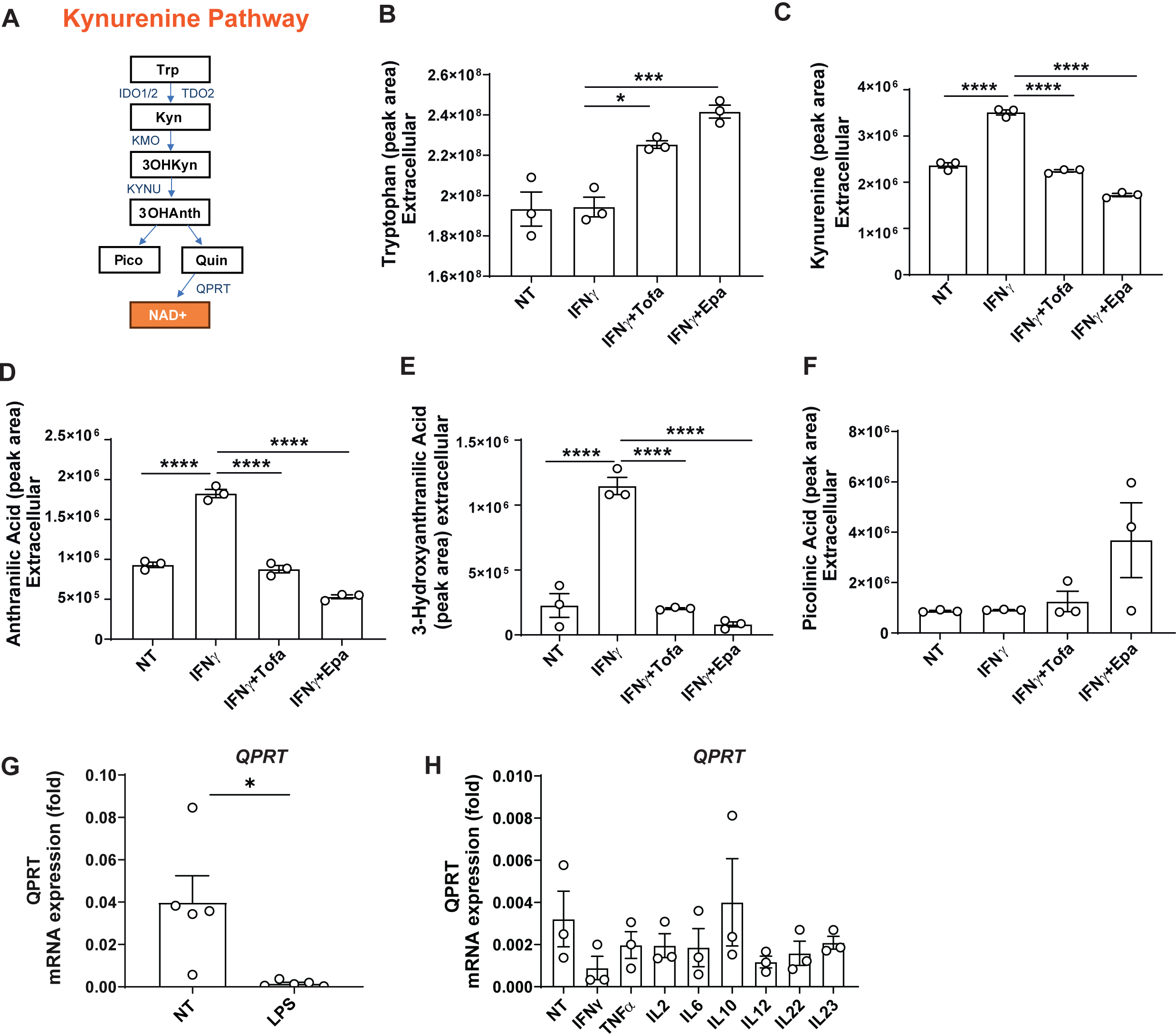
Quin accumulation following IFNγ-induced Trp catabolism is prevented by JAK inhibition. (A) Overview of the Kynurenine Pathway (KP). Trp: Tryptophan, Kyn: Kynurenine, 3OHKyn: 3-Hydroxy-kynurenine, 3OHAnth: 3-Hydroxyanthranilic acid, Pico: Picolinic acid, Quin: Quinolinic acid, NAD^+^: Nicotinamide adenine dinucleotide, IDO1/2: Indoleamine 2,3- dioxygenase 1/2, TDO: Tryptophan-2,3-dioxygenase, KMO: Kynurenine-3-monooxygenase, KYNU: Kynureninase, QPRT: Quinolinate phosphorybosyltransferase. (B-F) Human PBMC were incubated with IFNγ (10 ng/ml) ± tofacitinib (Tofa, 1 µM) or epacadostat (Epa, 5 µM) for 24 hours. Depicted metabolites were detected by LC-MS in cell culture supernatants (intracellular proportions of this experiment are found in Fig. 7B-G; n = 3 technical replicates, representative of n = 3 individual experiments). Metabolite abundance was normalized to cell numbers. (G) Human PBMC were treated with LPS (100 ng/ml) for 20 hours. Relative gene expression was assessed by RT-qPCR (n = 2 technical replicates, representative of n = 5 individual experiments; part of the gene expression analysis found in Fig. 6A). (H) Relative gene expression in human PBMC was detected after 24 hours of stimulation with 10 ng/ml of the named pro- or anti- inflammatory cytokines by RT-qPCR (n = 2 technical replicates, representative of n = 4 individual experiments; this gene expression analysis was performed within the experimental set-up of Fig. 6C,D). ß2-Mikroglobulin was used as a housekeeping gene. All data are presented as mean ± SEM. Statistical analysis was performed with one-way ANOVA (B-F,H) and an unpaired Student’s t-test (G). ∗*P* < 0.05; ∗∗∗*P* < 0.001; ∗∗∗∗P < 0.0001.

**Figure S9:**
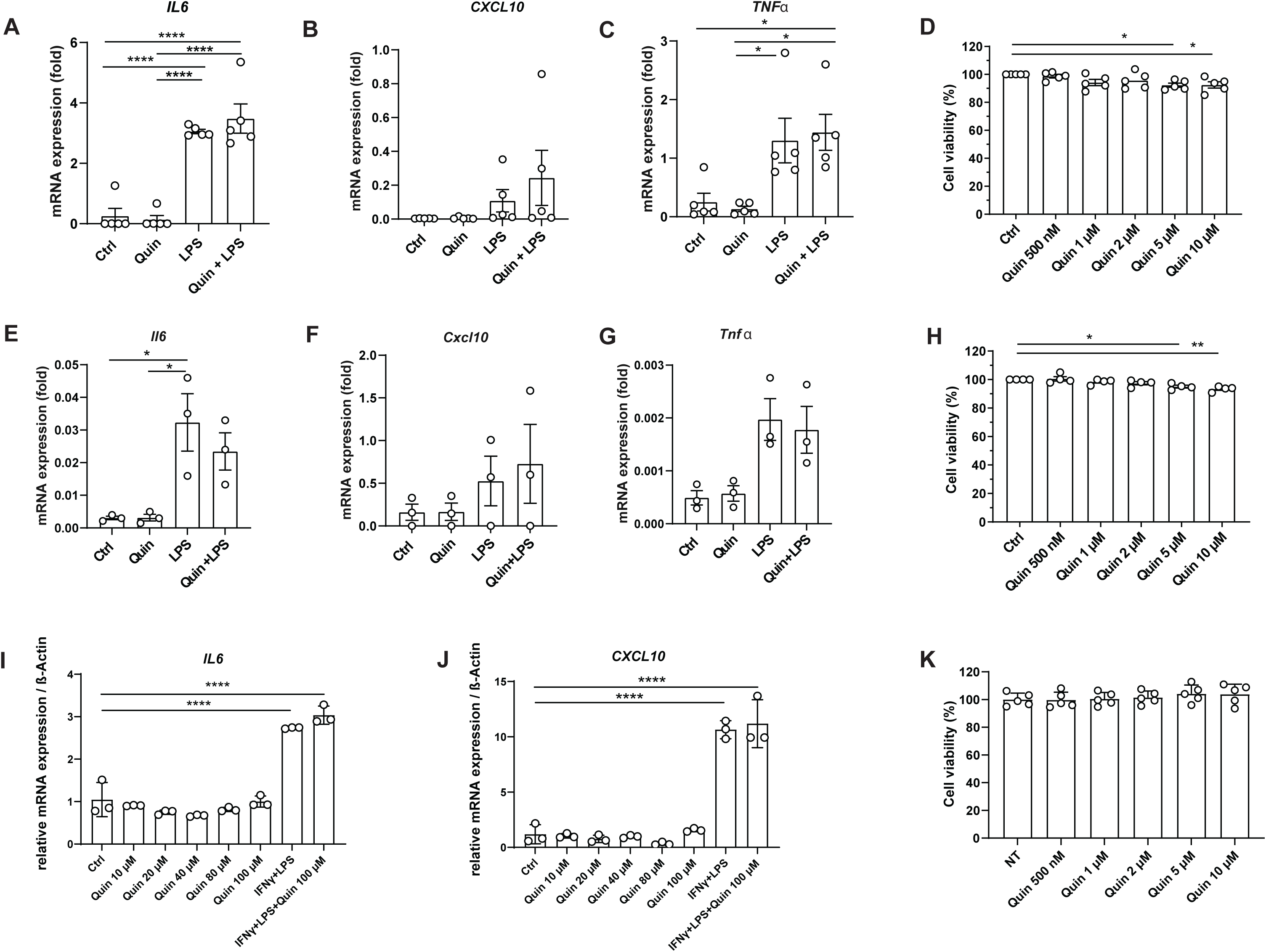
Extracellular Quin does not influence the pro-inflammatory cytokine expression. Human PBMC (A-C) and murine ModeK cells (E-G) were pre-incubated with quinolinic acid (10 µM) for 24 hours and then stimulated with LPS for 4 hours (for each cell type, n = 2 technical replicates, representative of n = 5 (PBMC) and n = 3-4 (ModeK) individual experiments were conducted). Gene expression was assessed by RT-qPCR, ß-Mikroglobulin served as a housekeeper. Human fibroblasts (I,J) were incubated with Quinolinic acid (Quin) for 24 hours at the indicated concentrations, IFNy (10 ng/ml for 24 hours) and LPS (1 µg/ml for 2 hours). Gene expression was assessed by RT-qPCR, ß-Actin served as a housekeeper (n = 3 technical replicates, representative of n = 3 individual experiments). For MTS assays, PBMC (D), ModeK cells (H) and fibroblasts (K) were incubated with the indicated concentrations of Quin for 24 hours. Relative cell viability was quantified in comparison to the viability of untreated cells (for each cell type, n = 3 technical replicates, representative of at least n = 3 individual experiments). Statistical analysis with one-way ANOVA. All data are presented as mean ± SEM. ∗P < 0.05; ∗∗P < 0.01; ∗∗∗∗P < 0.0001.

## Supplementary tables

**Supplementary Table 1.** Overview of the first longitudinal cohort, where HPLC Trp values were obtained and plotted over time. Age is calculated in years.

|  | Overall |
| --- | --- |
| No. patients | 180 |
| No. observations | 812 |
| Diagnosis (%) |  |
| CD | 99 (55.0) |
| UC | 81 (45.0) |
| Therapy persistence 12m (%) |  |
| 12m persistence | 120 (66.7) |
| Non-persistence | 60 (33.3) |
| Baseline tryptophan (mean (SD)) | 49.41 (18.37) |
| Sex = Female/Male (%) | 85/95 (47.2/52.8) |
| Age at baseline (mean (SD)) | 39.67 (13.55) |
| BMI (mean (SD)) | 25.64 (6.16) |
| Administered biologic (%) |  |
| Adalimumab | 20 (11.1) |
| Golimumab | 1 (0.6) |
| Infliximab | 65 (36.1) |
| Risankizumab | 2 (1.1) |
| Tocilizumab | 3 (1.7) |
| Tofacitinib | 10 (5.6) |
| Ustekinumab | 30 (16.7) |
| Vedolizumab | 49 (27.2) |
| Observations by time in weeks (%) |  |
| 0 | 156 (19.2) |
| 2 | 143 (17.6) |
| 6 | 158 (19.5) |
| 14 | 160 (19.7) |
| 26 | 92 (11.3) |
| 52 | 103 (12.6) |

**Supplementary Table 2.** Overview of the cohort for which Trp derivatives were examined. Age is given in years.

|  | Overall |
| --- | --- |
| No. patients | 134 |
| No. observations | 343 |
| Sex = Female/Male (%) | 63/71 (47.0/53.0) |
| Age at baseline (mean (SD)) | 38.38 (15.56) |
| Diagnosis (%) |  |
| CD | 52 (38.8) |
| UC | 82 (61.2) |
| Remission (%) |  |
| Non-remission | 57 (62.6) |
| Remission | 34 (37.4) |
| Administered biologic (%) |  |
| Adalimumab | 6 (4.5) |
| Filgotinib | 7 (5.2) |
| Golimumab | 1 (0.7) |
| Infliximab | 54 (40.3) |
| Ozanimod | 2 (1.5) |
| Tofacitinib | 6 (4.5) |
| Ustekinumab | 27 (20.1) |
| Vedolizumab | 31 (23.1) |
| Observations by time in weeks (%) |  |
| 0 | 133 (38.8) |
| 2 | 106 (30.9) |
| 14 | 104 (30.3) |

**Supplementary Table 3.** An overview of the cohort of IBD patients of which mucosal intestinal biopsies and whole blood were sampled for transcriptomics and metabolomics between week 0 and week 14 after initiation of a new biologic therapy. Age is given in years.

|  | Overall |
| --- | --- |
| No. patients | 62 |
| No. observations | 586 |
| Sex = Female/Male (%) | (61.3/38.7) |
| Age at baseline (mean (SD)) | 36.39 |
| Diagnosis (%) |  |
| CD | 32 (51.6) |
| UC | 30 (48.4) |
| Remission (%) |  |
| Non-remission | 28 (45.2) |
| Remission | 23 (37.1) |
| Non-inflamed | 11 (17.7) |
| Administered biologic (%) |  |
| Vedolizumab | 21 (33.9) |
| Infliximab | 22 (35.5) |
| Olamkicept | 16 (25.8) |
| Tocilizumab | 3 (4.8) |
| Observations by time in days (%) |  |
| 0 | 117 (20.0) |
| 0.17 (4-hours) | 62 (10.6) |
| 1 | 63 (10.8) |
| 3 | 29 (4.9) |
| 14 | 112 (18.9) |
| 42 | 109 (18.6) |
| 98 | 94 (16.0) |

## Methods

### Patient consent

Clinical studies conducted at the University Hospital Schleswig-Holstein, Campus Kiel, were approved by the ethics committee of the Christian Albrecht University of Kiel (D 490/20, D 489/20, A 124/14 and AZ 156/03-2/13) in accordance with the Declaration of Helsinki and all enrolled subjects provided written informed consent. All the following experimental and analytical procedures were carried out in accordance with relevant guidelines and regulations. Diagnoses, demographics, baseline characteristics, medication, numbers of observations and outcome (remission / non-remission at week 14 / therapy adherence at week 52) were recorded (Supplementary Tables 1-3).

### Statistical analysis for *in-vitro* data and DSS colitis

Statistical tests were selected with respect to data distribution and variance characteristics and non-parametric or parametric analyses were applied as appropriate: For *in-vitro* experiments, unpaired Student’s t-tests were employed when comparing two groups, while for all comparisons between multiple groups, one-way ANOVA with Šidák correction was used. For animal experiments, comparisons of two groups were conducted with Mann-Whitney U testing, whereas multiple groups were compared with Kruskal-Wallis tests and Dunn’s correction. We considered a threshold of 0.05 statistical significance of raw and adjusted p-values. The applied tests have been reported in the figure legends. Statistical tests were performed with GraphPad PRISM software 10 (GraphPad Software, La Jolla, CA, USA).

### Data availability

Available upon request.

### High performance liquid chromatography (HPLC)

Serum levels of tryptophan were determined as part of standard clinical laboratory assessments using a high-performance liquid chromatography (HPLC) kit, certified under In Vitro Diagnostic Conformité Européenne (CE) standards. The lower limits of detection and quantification were established according to the DIN 32645 guidelines.

### Targeted mass spectrometry for tryptophan derivative assessment in human serum

Samples were shipped frozen to Biocrates Life Sciences AG in Innsbruck, Austria, where they were kept frozen at −80C until they were measured using their ‘Tryptophan Metabolism Assay’. Briefly, protein was precipitated using a mixture of methanol and acetonitrile, and polar metabolites were added as internal standards. Samples were vortexed and centrifuged, with the supernatant dried and resuspended in water for measurement. Samples were split and measured with ultra high-performance liquid chromatography mass spectrometry (UHPLC-MS/MS) with multiple reaction monitoring in positive mode using SCIEX API 5500 QTRAP® (AB SCIEX, Darmstadt, Germany) and electrospray ionization. The second half of the measurements were performed on the same machine after a phenyl isothiocyanate derivitization step. All analytes were quantified using an external 7-point calibration.

Data were filtered according to the 80% rule ^63^: Only metabolites with 80% or greater non-missing values were retained for analysis. Metabolites with concentrations below the limit of quantitation and detection were considered missing. Following filtering, 3-OH-anthranilic acid, quinaldic acid, xanthureninc acid and 3-IPA had missing values at the following frequencies: 1.2%, 2.9%, 2.3%, and 6.1%, respectively. These values were imputed as half of each metabolite’s minimum observed value. Metabolites were log_10_ transformed and auto-scaled prior to statistical analysis.

We made use of the MetaboINDICATOR^TM^, as supplied by Biocrates, to further reveal the biological significance of the metabolic changes observed through the kit. In addition to the pre-calculated ratios provided, we expanded our panel to include two ratios based on the previous work of Michaudel et al ^19^. These two ratios, Quin:sumIDO and Quin:3OHAnth, were increased in their study during disease flares. Quin:sumIDO was calculated with in the following manner: Quin / (3-OH-anthranilic acid + 3-OH-kynurenine + kynurenic acid + kynurenine + picolinic acid + quinolinic acid + xanthurenic acid). In addition, we included the ratio of Quin:Trp as a measure of overall Quin synthesis rates relative to the entire Trp pool.

### Statistical analysis for the analysis of serum Trp metabolites

We included the following potential confounding variables for our linear mixed models used to assess changes to Trp over the first 52 weeks of biologic therapy administration: biologic therapy administered, diagnosis (UC/CD), body mass index, age at baseline, recruitment cohort, and sex assigned at birth (hereafter referred to as sex). The linear mixed models for the targeted metabolomics assay included the following additional covariates: sex, age body mass index, biologic status (i.e., whether a patient was taking a biologic therapy at the time of the observation), and, where appropriate, the diagnosis. Spearman correlations were performed using observations from week 14, where, owing to treatment, we expected the widest variance in disease activity. All analyses were performed using R version 4.4.0.

### Animal use and care

Animal experiments were performed at Pennsylvania State University, Pennsylvania, US, and approved by the Institutional Animal Care and Use Committee. Male 8-12 week old C57BL/6J mice were purchased from The Jackson Laboratory and received laboratory diet 5010 and water ad libitum while they acclimated for at least 7 days in a specific pathogen-free facility. Animals were exposed to a 12-hour-light-dark cycle (7 a.m. – 7 p.m.) before experimental use.

### Acute DSS-induced colitis

For DSS treatment, mice were single-housed and supplied with 2.5 % of DSS (MP Biomedicals) dissolved in autoclaved drinking water for 5 days, followed by treatment with regular drinking water. DSS-containing water was exchanged every other day. The disease activity index was assessed for each mouse daily by assessing weight loss in comparison to initial body weight, stool consistency, rectal bleeding, spontaneous behavior and activity as well as fur appearance. Consumption of drinking water was monitored daily.

### Chemicals and reagents for LC-MS/MS analysis of human tryptophan metabolites

Tryptophan (T0254), Kynurenine (K8625), Kynurenic acid (K3375), Quinaldic acid (160660), Anthranilic acid (10680), 3-Hydroxykynurenine (H1771), Picolinic acid (P42800), 5- Hydroxytryptophan (H9772), Serotonin (14927), 3-Indolepropionic acid (220027-1G), Nicotinamide (N0636), Xanthurenic acid (D120804), Nicotinic acid (72309) and 3- Hydroxyanthranilic acid (148776) and 3-Indoleacetic acid (45533) were purchased from Sigma Aldrich. Quinolinic acid (A11414) was purchased from ThermoFisher.

### Sample preparation of human PBMC for mass spectrometry

Extraction was carried out by incubating the cells in 1 ml of 100 % methanol for 20 minutes on a shaker at 4 °C. Supernatants were collected and concentrated overnight at −4 °C, using a Refrigerated CenrtiVap (Labconco, Missouri). Residues were reconstituted in 100 µl of 10 % Methanol, transferred to LC-vials and immediately measured by LC-MS/MS.

### Measurement of tryptophan metabolites in human PBMC by LC-MS/MS

10µL of each 4° C cold sample were injected in the LC-MS system. Chromatographic separation was performed with a Hypersil Gold C18 1.9 µm (150 x 2.1 mm) column (ThermoFisher Scientific) at 40° C using mobile phases A (0.1% formic acid) and B (Acetonitrile + 0.1% formic acid). Nitrogen was used for fragmentation and collision energy varied from 10 to 45 eV. Produced fragments were chosen for analyte quantification by Multiple Reaction Monitoring (m/z parents: m/z fragments). The analysis was performed on a HPLC (ExionLC, AB Sciex) coupled to a triple quadrupole mass spectrometer model QTRAP 5500+ (AB Sciex). The operation was done in positive electrospray ionization mode (ESI) in one single run. The area of the intensity vs. retention time was integrated for each analyte. Data were analysed with the program *Analyst* (version 1.7).

### Sample preparation for murine serum and tissues for mass spectrometry

Serum was thawed on ice before adding −80° C 100 % 80:20 methanol:water with a volume of 13.5 mL solvent / 1mL serum. Samples were vortexed, incubated on dry ice for 10 minutes and centrifuged at 4 °C, 16,000 g for 25 minutes. Supernatants were used for LC-MS analysis. Frozen tissues were weighed and grounded with liquid nitrogen in a cryomill (Retsch) at 25 Hz for 45 seconds. A mix of 40:40:20 acetonitrile:methanol:water with a volume of 40 μL solvent / 1mg of tissue was added, before samples were vortexed for 15 seconds, and incubated on ice for 10 minutes. Tissue samples were then centrifuged at 4 °C and 16,000 g for 30 minutes. Supernatants were transferred to new Eppendorf tubes and then centrifuged at 16,000 g for 25 minutes to remove residual debris. This step was repeated before supernatants were transferred to MS vials and loaded to the LC-MS.

### Metabolite measurement of murine serum and tissues for mass spectrometry

Extracts were analysed within 24 hours after preparation by liquid chromatography coupled to a mass spectrometer (LC-MS). The LC-MS method involved hydrophilic interaction chromatography (HILIC) coupled to the Q Exactive PLUS mass spectrometer (ThermoFisher Scientific) ^64^. The LC separation was performed on a XBridge BEH Amide column (150 mm 3 2.1 mm, 2.5 mm particle size, Waters, Milford, MA). Solvent A was 95%:5% H2O: acetonitrile with 20 mM ammonium bicarbonate, and solvent B was acetonitrile. The gradient was 0 min, 85% B; 2 min, 85% B; 3 min, 80% B; 5 min, 80% B; 6 min, 75% B; 7 min, 75% B; 8 min, 70% B; 9 min, 70% B; 10 min, 50% B; 12 min, 50% B; 13 min, 25% B; 16 min, 25% B; 18 min, 0% B; 23 min, 0% B; 24 min, 85% B; 30 min, 85% B. Other LC parameters were: flow rate 150 ml/min, column temperature 25° C, injection volume 10 μL and autosampler temperature was 5° C. The MS was operated in both negative and positive ion mode for the detection of metabolites. Other MS parameters used were: resolution of 140,000 at m/z 200, automatic gain control (AGC) target at 3e6, maximum injection time of 30 ms and scan range of m/z 75-1000. Raw LC-MS data were converted to mzXML format using the command line ‘msconvert’ utility ^65^. Data were analysed via the MAVEN software.

### Integrative transcriptomics and metabolomics analysis

Whole blood and biopsies were obtained within two longitudinal IBD intervention cohorts sampled in northern Germany (n = 62 IBD, n = 32 UC, n = 30 CD) over a period of 14 weeks at the indicated timepoints. Patients were treatment-naïve for biologic treatment and were introduced to either anti-TNFα (n = 22), anti-α4β7-Integrin (n = 21), anti-IL6-trans (n = 16) or anti-IL6-R (n = 3). Patients were clinically and endoscopically monitored. N = 120 biopsies and n = 123 whole bloods samples were used for RNA sequencing, whereas n = 128 serum samples were used for metabolomics using the Biocrates MxP Quant 500 kit (Biocrates Life Sciences AG, Innsbruck, Austria) according to manufacturer’s instructions and as described in detail previously ^22^. Differential gene expression analysis was performed using variance stabilized read counts (DESeq2) and afterwards applying linear mixed models (lme4) by using the following formula: vst ∼ Sex * <HBI/Mayo|kyn/trp> + (1|PatientID). Enrichment analyses were performed by means of gene set enrichments (GSEA) or hypergeometric tests for overrepresentation (ClusterProfiler). For GSEA, a ranking vector was created by summing the t-values for the main effect of HBI/Mayo or serum Kyn:Trp ratio and the interaction with sex. All analyses were performed in R and all p-values were adjusted with the Benjamini- Hochberger correction.

### Transcriptomics & metabolic modelling

Employing the RNA sequencing used within the above-mentioned integrative analysis, the human metabolic model (recon3D) was utilized to extract the Trp degradation pathway. The expression values (as TPM) were mapped to Trp pathways applying the gene-protein-reaction relations of recon3D (github.com/Porthmeus/CORPSE) for these reactions to estimate reaction activity scores (RAS). RAS were than associated with HBI/Mayo in linear mixed models with the following formula: rxnExpr ∼ Sex * <HBI/Mayo> + (1|PatientID). All p-values were adjusted with the Benjamini-Hochberger correction. For missing HBI/Mayo scores, we used linear modeling and imputed values according to the given time trajectory of the individual patient during the time course assuming a logarithmic decline in HBI/Mayo scores over time.

### *IDO1* and *QPRT* gene expression analysis in human intestinal biopsies

Published microarray and RNA-seq data sets were employed to assess normalized *IDO1* and *QPRT* mucosal expression from biopsy samples of UC patients. Endoscopic improvement was defined as a Mayo endoscopic subscore ≤1, as described previously ^62^. Biopsies were collected within different longitudinal IBD treatment cohorts: **i)** Highly refractory UC patients pre-exposed to infliximab and vedolizumab were treated with Tofacitinib ^62^. Biopsies from 4 responders and 11 non-responders were included. **ii)** A prospective observational study of IBD subjects with active colonic disease who received vedolizumab or infliximab for induction of remission ^24,25^. 32 longitudinal biopsy samples of patients receiving vedolizumab were included (baseline = 9; week 2 = 7; week 6 = 8; week 14 = 8), whereas for patients treated with infliximab, 28 samples were used (baseline = 8; week 2 =7; week 6 = 7; week 14 = 6). RNA sequencing of whole colonic biopsies was performed, and raw gene expression counts were normalized using DESeq2 R package ^66^, version 1.40.2. RNA sequencing of whole colonic biopsies was performed, and raw gene expression counts were normalized using DESeq2 R package ^66^, version 1.40.2.

### Correlation of mucosal gene expression with endoscopic and histologic disease severity as well as ISG scores

Mucosal expression of *IDO1* and *QPRT* were extracted from the publicly available transcriptomics datasets GSE73661 (microarray of UC patients) and GSE109142 (RNA sequencing TPM counts of pediatric ulcerative colitis patients) that were downloaded from the Gene Expression Omnibus (GEO) repository and processed using R version 4.2.2. *IDO1* and *QPRT* gene expression levels were correlated with i) the Mayo endoscopic subscore (MES) for 166 biopsies from 67 UC patients (treated with either infliximab or vedolizumab) and 12 samples from 12 healthy donors (GSE73661 ^30^), ii) the histology severity score (HSS) of 206 new-onset UC patients and 20 healthy controls (GSE109142 ^29^). For correlation of *IDO1* and *QPRT* expressions with pro-inflammatory signaling processes, we calculated IFNγ, TNFα and IL-23 scores using the *R* package *singscore* ^67–70^ and the IFNγ geneset composed of *IFITM1*, *MX1*, *OAS3*, *IFIT1*, *IFI44L* and *IFI16* ^71^ as well as the TNF and IL-23 genesets described in Martínez et al. ^70^.

### Single cell sequencing analysis of *IDO1* and *QPRT* expression across intestinal cell types

Single cell RNAseq analysis was conducted with intestinal tissues obtained from 13 fresh full-thickness CD resections (noninvolved or inflamed nonstrictured segments) as well as from 7 normal non-CD bowel segments as described previously ^36^. Expression of *QPRT*, *IDO1* and *IDO2* genes was evaluated in the single cell RNAseq dataset with a Shiny app created using the R package ShinyCell (Ouyang JF. ShinyCell: Shiny Interactive Web Apps for Single-Cell Data. R package version 2.1.0, https://github.com/SGDDNB/ShinyCell).

### Supplemental Materials and Methods

#### RNA isolation, cDNA synthesis and gene expression analysis

Using the RNeasy Kit (Qiagen), mRNA was isolated from different cell types. cDNA was synthesized using Monarch Genomic DNA Purification Kit (New England Biolabs) according to the manufacturer’s protocol. To examine gene expression, cDNA samples were subjected to quantitative reverse transcription polymerase chain reaction using TaqMan assays purchased from Integrated DNA Technologies (for TaqMan IDs and RefSeq numbers, see Supplementary Table 4). Reactions were carried out on the Applied Biosystems 7900HT Fast Real-Time PCR System (Applied Biosystems) and relative transcription levels were determined utilizing ß2-Microglobulin (B2M) or ß-Actin as housekeeper.

**Supplementary Table 4:** Primers used for gene expression analysis.

| Gene name | Species | Taqman ID | RefSeq Number |
| --- | --- | --- | --- |
| IDO1 | Human | Hs.PT.58.924731 | NM_002164 |
| IDO2 | Human | Hs.PT.58.3013208 | NM_194294 |
| IL10 | Human | Hs.PT.58.2807216 | NM_000572(1) |
| IL12B | Human | Hs.PT.58.2925830 | NM_002187(1) |
| IL2 | Human | Hs.PT.58.1142676 | NM_000586(1) |
| IL22 | Human | Hs.PT.58.1865033 | NM_020525(1) |
| IL23A | Human | Hs.PT.58.38730006.g | NM_016584(1) |
| IL6 | Human | Hs.PT.58.40226675 | NM_000600(1) |
| CXCL10 | Human | Hs.PT.58.3790956g | NM_001565(1) |
| JAK2 | Human | Hs.PT.58.25272310 | NM_004972(1) |
| NMNAT3 | Human | Hs.PT.58.18999775 | NM_001200047 |
| QPRT | Human | Hs.PT.58.142969 | NM_014298 |
| TDO2 | Human | Hs.PT.58.3092178 | NM_005651 |
| TNFα | Human | Hs.PT.58.45380900 | NM_000594(1) |
| β-Actin | Human | Hs.PT.39a.22214847 | NM_001101(1) |
| β2-Mikroglobulin | Human | Hs.PT.58v.18759587 | NM_004048 |
| Cxcl10 | Murine | Mm.PT.58.43575827 | NM_021274(1) |
| IL6 | Murine | Mm.PT.58.10005566 | NM_031168(1) |
| TNF $\alpha$ | Murine | Mm.PT.58.12575861 | NM_013693(1) |
| β-Actin | Murine | Mm.PT.39a.22214843.g | NM_007393(1) |

#### Immunoblot analysis

Cells were lysed using RIPA buffer with 1 % Halt™ Protease and Phosphatase Inhibitor Cocktail (ThermoFisher Scientific). Lysates were centrifuged at 16,000 g for 15 minutes at 4 °C, diluted with PBS and 5x Laemmli buffer, and heated at 95 °C for 10 minutes. Afterwards, equal amounts of protein lysates containing Laemmli buffer were electrophoresed on 12 % polyacrylamide gels under standard SDS-PAGE conditions before being transferred onto polyvinylidene fluoride membranes (GE Healthcare). Protein-loaded membranes were blocked with 5 % milk in tris buffered saline and Tween 20 (TBST) before being incubated with the primary antibody at 4 °C overnight followed by incubation with a horseradish peroxidase– conjugated secondary antibody for 1 hour at indicated concentrations (Supplementary Table 5). Proteins were detected using the Amersham ECL Prime Western Blot Detection Reagent (GE Healthcare).

**Supplementary Table 5:** Antibodies used for western blot analysis.

| Primary Antibody | Origin | Dilution | Company | Article number |
| --- | --- | --- | --- | --- |
| Phospho-STAT1 (Tyr701) (58D6) | Rabbit | 1:1000 | Cell Signaling Technology | 9167 |
| Phospho-STAT3 (Tyr705) (D3A7) | Rabbit | 1:1000 | Cell Signaling Technology | 9145 |
| STAT1 | Rabbit | 1:1000 | Cell Signaling Technology | 9172 |
| STAT3 (D3Z2G) | Rabbit | 1:1000 | Cell Signaling Technology | 12640 |
| β-Actin | Mouse | 1:5000 | Sigma Aldrich | A5441 |
| IDO (D5J4E<sup>™</sup>) | Rabbit | 1:1000 | Cell Signaling Technology | 86630 |
| QPRT | Rabbit | 1:1000 | Proteintech | 25174-1-AP |

| Secondary Antibody | Origin | Dilution | Company | Article number |
| --- | --- | --- | --- | --- |
| Amersham ECL Rabbit IgG, HRP-linked whole Ab (from donkey) | Rabbit | 1:2000 | Cytiva | NA934 |
| Amersham ECL Mouse IgG, HRP-linked whole Ab (from sheep) | Mouse | 1:2000 | Cytiva | NA931 |

#### NAD(H) detection by GloAssay

To measure the total intracellular NAD^+^ and NADH levels, we used the NAD/NADH-Glo^TM^ Assay (G9071, Promega) following the instructions provided by the manufacturer. Cells were treated with different stimulants. Afterwards, cells were transferred to a white-walled, clear flatbottom 96-well plate and mixed with an equal amount of detection reagent to lyse the cells. The plates were maintained at room temperature for 60 minutes, before luminescence was measured using a Tecan microplate reader. The NAD(H) ratio was determined by comparing the luminescence of stimulated cells with that of untreated cells.

#### MTS assay

Viability of different cell lines cultivated on 96-well plates was assessed by measuring 3-(4,5-dimethylthiazol-2-yl)-5-(3-carboxymethoxyphenyl)-2-(4-sulfophenyl)-2H-tetrazolium (MTS) incorporation using the CellTiter 96 AQueous One Solution Cell Proliferation Assay (Promega) according to the manufacturer’s protocol.

#### Transfection of cells with siRNA (small interfering RNA)

For *in-vitro* transfection, Lipofectamine RNAiMAX was used according to the manufacturers’ protocol. SiRNA against *QPRT* (#GS67375) was derived from Qiagen and used at a concentration of 10 µM.

#### Human Peripheral Blood Mononuclear Cells

Human PBMC (Peripheral Blood Mononuclear Cells) were isolated from human buffy coats obtained from healthy adult volunteers following the guidelines of the University Hospital of Schleswig-Holstein. 9 ml of blood were diluted with an equal amount of PBS and transferred into a 50 ml centrifuge tube pre-filled with 9 ml of Histopaque-1077 (Sigma Aldrich). After centrifugation at 800 g for 20 minutes at room temperature, the collected layer of PBMC was washed by adding PBS and centrifuged again at 400 g for 7 minutes at 4 °C. After conducting the washing step twice, PBMC were cultured in RPMI (Invitrogen) containing 10% FBS (SERANA).

#### Human fibroblasts

The human colon CCD-18Co myofibroblast cell line (ATCC CRL-1459) was cultivated at 37°C with DMEM medium (Gibco), enriched with 10% FCS (Merck Millipore) and supplemented with 1% NEAA 100x (Gibco) and 1% penicillin streptomycin (Gibco). Medium was exchanged every other day and cells were sub-cultured at 80-85% confluence.

#### Murine enterocytes

ModeK cells were immortalized by employing the SV40 Virus (simian vacuolating virus). Cells were cultivated at 37 °C with DMEM GlutaMAX™ medium (Gibco,) with 10 % FCS (Merck Millipore). Medium was exchanged every other day.

#### Stimulants for *in-vitro* experiments

As stimulants for PBMC, ModeK cells and fibroblast, the stimulants (company, article number) listed in Supplementary Table 6 were used.

**Supplementary Table 6:** Stimulants used for cell culture.

| Name | Company | Article number |
| --- | --- | --- |
| Tofacitinib | Sigma Aldrich | PZ0017 |
| Upadacitinib | Selleck Chemicals | S8162 |
| Filgotinib | Selleck Chemicals | S7605 |
| Epacadostat | Selleckchem | S7910 |
| LPS (Lipopolysaccharides, Escherichia coli O111:B4) | Sigma Aldrich | L2630 |
| 2,3-Pyridinedicarboxylic acid | Sigma Aldrich | P63204 |
| Quinolinic Acid | ThermoFisher Scientific | 11440253 |
| rh IFN-gamma | Gibco | PHC4031 |
| rh IFN-gamma | Immunotools | 11343534 |
| rh IL-10 | immunotools | 11340102 |
| rh IL-22 | Immunotools | 11340222 |
| rh IL-23 | Immunotools | 11340232 |
| rh IL-6 | Immunotools | 11340060 |
| rh IL-12 | Immunotools | 11349122 |
| rh TNF-alpha | Immunotools | 11343013 |
| rh IL-2 | Immunotools | 11340023 |

## Notes

**Grant support** This work was supported by the BMBF iTREAT project (P.R., C.K.), DFG Cluster of excellence (ExC2167) “Precision medicine in chronic inflammation” RTF III, RTF-VIII and TI-1, the DFG CRC 1182 C2 (P.R.), the EU project miGut-Health (P.R.), the EKFS research grant #2019_A09 and EKFS Clinician Scientist Professorship (K.A., 2020_EKCS.11), the BMBF (eMED Juniorverbund “Try-IBD” 01ZX1915A, 01ZX2215, K.A., D.H.), the DFG RU5042 (P.R., K.A., C.K.), the Joachim Herz Stiftung (K.A.), NIH Grant T32GM108563 (A.I.A.), the Howard Hughes Medical Institute Hanna H. Gray Fellows Program Faculty Phase (Grant# GT15655, M.R.M), the Burroughs Welcome Fund PDEP Transition to Faculty (Grant# 1022604, M.R.M), the ECCO Multiyear Research Grant 2021 (K.A., B.V). and the Clinical Research Fund (KOOR) at the University Hospitals Leuven and the Research Council at the KU Leuven (B.V.). This project has furthermore received funding from the Innovative Medicines Initiative 2 Joint Undertaking (JU) under grant agreement No 853995 (ImmUniverse). The JU receives support from the European Union’s Horizon 2020 research and innovation programme and EFPIA.

### Competing Interest Statement

The authors have declared no competing interest.

### Funding Statement

This work was supported by the BMBF iTREAT project (P.R., C.K.), DFG Cluster of excellence
(ExC2167) Precision medicine in chronic inflammation RTF III, RTF-VIII and TI-1, the DFG
CRC 1182 C2 (P.R.), the EU project miGut-Health (P.R.), the EKFS research grant #2019_A09
and EKFS Clinician Scientist Professorship (K.A., 2020_EKCS.11), the BMBF (eMED
Juniorverbund Try-IBD 01ZX1915A, 01ZX2215, K.A., D.H.), the DFG RU5042 (P.R., K.A.,
C.K.), the Joachim Herz Stiftung (K.A.), NIH Grant T32GM108563 (A.I.A.), the Howard
Hughes Medical Institute Hanna H. Gray Fellows Program Faculty Phase (Grant# GT15655,
M.R.M), the Burroughs Welcome Fund PDEP Transition to Faculty (Grant# 1022604, M.R.M),
the ECCO Multiyear Research Grant 2021 (K.A., B.V). and the Clinical Research Fund (KOOR)
at the University Hospitals Leuven and the Research Council at the KU Leuven (B.V.). This
project has furthermore received funding from the Innovative Medicines Initiative 2 Joint
Undertaking (JU) under grant agreement No 853995 (ImmUniverse). The JU receives support
from the European Union's Horizon 2020 research and innovation programme and EFPIA.

### Author Declarations

Clinical studies conducted at the University Hospital Schleswig-Holstein, Campus Kiel, were approved by the ethics committee of the Christian Albrecht University of Kiel (D 490/20, D 489/20, A 124/14 and AZ 156/03-2/13) in accordance with the Declaration of Helsinki and all enrolled subjects provided written informed consent

